# Prediabetes and glycemic transitions as determinants of frailty and functional decline in adults aged 50 years and older: A longitudinal analysis from five multinational aging cohorts

**DOI:** 10.64898/2026.04.22.26351540

**Authors:** Andrea Malagon-Liceaga, Martín Roberto Basile-Alvarez, Carlos A. Fermín-Martínez, Diana Laura Ramírez-Rivera, Jerónimo Perezalonso-Espinosa, Juan Pablo Díaz-Sánchez, Sara Berenice García-González, Karime Berenice Carrillo-Herrera, Leslie Alitzel Cabrera-Quintana, Neftali Eduardo Antonio-Villa, Natalia Gomes Gonçalves, Carmen Garcia-Peña, Omar Yaxmehen Bello-Chavolla

## Abstract

**Background:** Prediabetes is highly prevalent in older adults and is characterized by heterogeneous clinical trajectories, including regression to normoglycemia and progression to diabetes. While prediabetes has been associated with impaired physical function and frailty, the longitudinal impact of both a single diagnosis and dynamic glycemic transitions on functional outcomes remains unclear. We aimed to evaluate associations between baseline prediabetes and glycemic transitions over time with trajectories of functional capacity and frailty in older adults.

**Methods:** We conducted a pooled analysis of harmonized data from five nationally representative longitudinal aging cohorts (MHAS, HRS, CHARLS, ELSA, CRELES) within the Gateway to Global Aging Data, including adults aged ≥50 years with ≥1 HbA1c measurements. Prediabetes was defined per ADA criteria (HbA1c 5.7-6.4%). Functional outcomes included activities of daily living (ADL), instrumental ADL (IADL), and frailty assessed using Fried phenotype, FRAIL scale, and a deficit-accumulation Frailty Index (FI). Mixed-effects Poisson models estimated incidence rate ratios (IRRs) for baseline prediabetes, while generalized estimating equations assessed time-varying glycemic status and transition trajectories. Models were adjusted for age, sex, cohort, and time-varying covariates, with sensitivity analyses including BMI, smoking, and alcohol intake.

**Findings:** Among 18,571 participants (median follow-up 13.6 years), baseline prediabetes was associated with increased progression of functional deficits and frailty compared with normoglycemia, including higher FI values and accelerated FI progression. Prediabetes was associated with higher incidence of ADL, IADL, and multimorbidity deficits from early follow-up, although time-dependent changes in incidence rates were not significant. In time-varying analyses (n=7,840), both prediabetes and diabetes were associated with higher incidence of functional deficits compared with normoglycemia, with diabetes showing the strongest effects across all outcomes. Diabetes was associated with greater FI burden and accelerated progression, whereas prediabetes showed a smaller increase, with attenuation over time. Among individuals with baseline prediabetes, regression to normoglycemia occurred in 20.8% and was associated with increased incidence of ADL and frailty deficits. In contrast, progression to diabetes occurred in 24.3%, and was associated with lower risk of incident ADL and Fried frailty deficits compared to stable prediabetes.

**Interpretation:** Prediabetes is associated with increased risk of functional decline, frailty, and deficit accumulation in older adults, independent of progression to diabetes. Regression to normoglycemia was associated with higher risk of functional deterioration. These findings suggest that prediabetes reflects a state of metabolic vulnerability linked to biological aging rather than solely a precursor to diabetes and highlights a need to reframe its clinical significance in older populations.

**Funding:** This research was supported by Instituto Nacional de Geriatría in Mexico.

**RESEARCH IN CONTEXT:** *Evidence before this study:* We searched PubMed up to April 1, 2026, using the terms (“prediabetes”) AND (“frailty” OR “functional decline” OR “activities of daily living” OR “mortality”) AND (“older adults” OR “aging” OR “longitudinal” OR “cohort”). Existing evidence from predominantly single-country and cross-sectional studies suggests that prediabetes is associated with increased risk of frailty, disability, and adverse health outcomes in older adults. However, findings have been inconsistent, particularly regarding incident versus prevalent outcomes and the role of competing risks in aging populations. Importantly, few studies have used harmonized multinational longitudinal data, incorporated repeated measures of glycemic status, or evaluated multiple complementary frailty constructs. The impact of dynamic glycemic transitions, including regression to normoglycemia and progression to diabetes, on functional trajectories remains poorly understood.

*Added value of this study:* This study leverages harmonized longitudinal data from five nationally representative aging cohorts from Mexico, England, China, Costa Rica and the United States to examine both baseline prediabetes and time-varying glycemic status in relation to trajectories of functional decline and frailty. By integrating multiple validated measures of functional capacity and frailty, and applying longitudinal modeling strategies, we provide a comprehensive assessment of the relationship between prediabetes and age-related outcomes. We further characterize glycemic transition trajectories and show that regression to normoglycemia among older adults with prediabetes is associated with increased risk of frailty, while progression to diabetes does not uniformly confer additional risk beyond stable prediabetes.

*Implications of all the available evidence:* Our results support a reconceptualization of prediabetes in older adults as a marker of systemic vulnerability rather than solely a precursor to diabetes. Glycemic status in later life appears to also reflect underlying physiological reserve, with dynamic changes potentially signaling deterioration rather than improvement. Our findings suggest a need to incorporate glycemic markers into geriatric risk stratification and to interpret their changes in the context of aging biology. Future research should focus on elucidating the mechanisms linking dysglycemia with frailty and functional decline, including the roles of sarcopenia, inflammation, and multimorbidity, and on evaluating interventions tailored to older populations across diverse global contexts.

## INTRODUCTION

Prediabetes is defined as intermediate state of dysglycemia between normoglycemia and diabetes, and is often considered primarily as a risk factor for the development of diabetes, while being accompanied by other associated metabolic disturbances^1^. Recent estimates indicate that impaired glucose tolerance affects ∼12.0% of adults globally, and impaired fasting glucose ∼9.2%^2^. In adults aged ≥60 years, prediabetes prevalence often exceeds 40-50% which has been attributed to age-related insulin resistance, β-cell dysfunction, mitochondrial changes, and reduced incretin effects^3–5^. Despite being common, the clinical course of prediabetes in older adults differs markedly from younger adults. Longitudinal studies show that in older adults with prediabetes regression to normoglycemia or death is far more common than progression to diabetes^3^. Rooney et al. previously reported, across various prediabetes definitions, reversion rates of 13-44% and mortality rates of 16-19%, versus progression to diabetes in only 8-9% of participants over 6 years^3^. Similarly, Shang et al. reported that over 12 years 41.3% of participants remained stable in prediabetes, 22.2% reverted to normoglycemia, 13.0% progressed to diabetes, and 23.4% died; importantly, weight loss and physical activity favoured reversion to normoglycemia^5^. However, the impact of these glycemic transitions on functional outcomes has not been evaluated.

Preservation of functional capacity is essential for autonomy in later life. Limitations in activities of daily living (ADL) and instrumental ADL (IADL) predict future disability and dependency, while frailty, reflecting reduced physiological reserve, strongly predicts adverse outcomes including hospitalization and mortality^6–9^. Prediabetes has been associated to impaired physical function, disability and frailty in older adults, with heterogeneous results across studies^10–12^. However, it is unclear whether the impact of this risk differs when considering a prediabetes diagnosis at a single time point compared to dynamic changes in glycemic status which also consider the possibility of regressing to normoglycemia or progressing to diabetes. Here, we aimed to examine longitudinal associations between a single prediabetes diagnosis at baseline and glycemic transitions over time with trajectories of functional capacity and frailty in a harmonized multinational sample of older adults.

## METHODS

### Data sources and participants

We analyzed longitudinal data from five nationally representative aging studies, harmonized through the Gateway to Global Aging Data platform (G2A, https://g2aging.org/). We included participants aged ≥50 years at baseline with at least one valid HbA1c measurement and two or more follow-up measurements from the Mexican Health and Aging Study (MHAS, Mexico)^13^, the Health and Retirement Study (HRS, United States)^14^, the China Health and Retirement Longitudinal Study (CHARLS, China)^15^, the English Longitudinal Study of Aging (ELSA, England)^16^, and the Costa Rican Longevity and Healthy Aging Study (CRELES, Costa Rica)^17^. Sampling designs for all cohorts are summarised in **Supplementary Methods**. For all participants, the baseline wave was defined as the first with available HbA1c measurement, and follow-up waves were harmonized into relative timepoints (t0 baseline, t1–t5 follow-up). Included waves and time mapping are detailed in **Supplementary Tables 1 and 2**. Analyses focused on a single diagnosis of prediabetes at baseline included all five G2A cohorts, whilst those focused on glycemic transitions which requires ≥2 HbA1c measures over time were limited to ELSA, HRS and CRELES.

### Prediabetes definition and glycemic transitions

HbA1c was harmonized as the sole glycemic biomarker across cohorts. Prediabetes was defined according to the American Diabetes Association (ADA) criteria as HbA1c 5.7-6.4% without prior self-reported diabetes, and diabetes as HbA1c ≥6.5% or prior self-reported diagnosis. We classified transitions from baseline prediabetes status into three categories as stable prediabetes (persistent prediabetes across waves), progression to diabetes (transition to self-reported diabetes or HbA1c ≥6.5% at any follow-up wave), or regression to normoglycemia (transition to HbA1c <5.7% at follow-up waves without subsequent diabetes). Diabetes was treated as a non-reversible state once progression from prediabetes occurred.

### Study outcomes

Functional capacity was assessed using Katz ADL (5 items: bathing, dressing, eating, transferring, toileting),) Barthel ADL (7 items, adding stairs and walking short distances), and instrumental ADL (IADL; 4 items: meal preparation, money management, shopping, medications). Frailty was measured with three constructs: (1) Fried phenotype defined using exhaustion via Center for Epidemiologic Studies Depression Scale (CES-D) items, weight loss ≥5–10% from the immediate previous wave, low grip strength defined below the lowest sex/BMI-adjusted quintile, slow gait defined as the slowest sex/height-adjusted quintile, and low vigorous activity; (2) FRAIL scale (fatigue, resistance/ambulation difficulties, ≥5 comorbidities, weight loss); (3) and a deficit-accumulation Frailty Index (FI) composed by 31 items across 9 domains: ADL, IADL, CES-D depressive symptoms, mobility, self-rated health/hearing/memory, polypharmacy, comorbidities, falls, low BMI; and 26 items in CRELES. All-cause mortality was registered from cohort records and time-to-event was estimated from interview date at baseline until the next interview date at each follow-up wave or recorded date of death. Detailed variable harmonisation, and FI specifications are provided in **Supplementary Tables 3 and 4**.

### Statistical analysis

#### Study population

Baseline characteristics were summarised using medians and interquartile range (IQR) for continuous variables and proportions for categorical variables. Mann-Whitney U-tests were conducted for bivariate analyses of continuous variables and chi-squared tests for categorical variables. Incidence rates new deficits were calculated amongst individuals without deficits at baseline as events per 1,000 person-years and were standardized by age and sex using the standard population ≥50 years across pooled G2A studies using the *epitools* R package. All analyses were performed using R software version 4.5.2.

#### Risk of functional and frailty deficits over time with a prediabetes diagnosis at baseline

Longitudinal associations between baseline prediabetes and the progression and incidence of functional deficits and frailty were evaluated using mixed effects generalized linear models with a Poisson log-link function to estimate incidence rate ratios (IRRs) and 95%CIs with robust standard errors. Repeated observations for covariates and outcomes across study waves were accounted for using participant-specific random effects, and an uncorrelated random slope for study wave (t0, t1, onwards). Outcomes were analyzed using several definitions to capture different aspects of deficit accumulation: (1) progression of deficits using the total count across waves, (2) incident deficits, and binary indicators of developing ≥1 or ≥2 incident deficits during follow-up. For incidence only participants without deficits at baseline were included, and the time variable was centered so that the first follow-up wave served as the reference. Models included baseline prediabetes as the primary exposure, a time variable, and a prediabetes by time interaction to assess differences in the rate of deficit accumulation.

#### Glycemic transitions over time

Amongst individuals with ≥2 HbA1c measurements over time, we examined transitions between glycemic states varying from normoglycemia, prediabetes and diabetes. Primary analyses included all participants from baseline to their last available HbA1c measurement during follow-up and additionally analyzing death as a censoring event. Secondary analyses were restricted to participants with HbA1c measurements between subsequent longitudinal waves, allowing assessment of transitions over shorter time periods.

#### Glycemic status and risk of functional deficits over time

To evaluate the longitudinal impact of time-varying glycemic status, analyses with participants categorized at each wave as normoglycemia, prediabetes, or diabetes. Diabetes was considered an absorbing state, such that individuals classified with diabetes at any wave remained as such in subsequent observations. Longitudinal associations between dynamic glycemic status with ADL, IADL, comorbidities and frailty were examined using generalized estimating equations (GEE) with Poisson log-link functions to estimate IRRs and 95%Cis with robust standard errors. Models incorporated an interaction term between glycemic status and years since baseline to assess rates of deficit accumulation. Correlation within individuals across repeated observations was accounted for using a first-order autoregressive correlation structure. For incidence analyses only participants without deficits at baseline were included.

#### Analyses of glycemic transition trajectories amongst participants with prediabetes

Among participants with prediabetes at baseline, glycemic trajectories were further classified as regression to normoglycemia, stable prediabetes, or progression to diabetes. Longitudinal associations with functional outcomes were assessed using GEE models with a Poisson log-link, with time since transition onset, and stable prediabetes serving as the reference category. These analyses were restricted to individuals with at least two consecutive observations following baseline and excluded periods prior to the first observed transition.

#### Frailty index trajectories

Longitudinal changes in the FI were modeled using both linear mixed-effects generalized linear models and GEE with Gaussian links. Mixed-effects linear models included random intercepts and uncorrelated slopes for follow-up time to account for individual-level heterogeneity in frailty trajectories for a prediabetes diagnosis at baseline. GEE models were implemented to assess the longitudinal impact of time-varying glycemic status on FI over time. Non-linear temporal effects were explored using restricted cubic splines.

#### Standardization, adjustment and weighting

All longitudinal models incorporated standardized survey response weights harmonized within each cohort and wave to account for differential sampling probabilities and attrition. Time-dependent offsets representing years since baseline were included in incidence models to estimate rates per person-year time. All models were adjusted for time-varying age, sex and study cohort. Additionally, to rule out an effect of prediabetes, regression to normoglycemia or progression to driven by lifestyle changes or by changes in BMI, we performed a sensitivity analysis with additional adjustment by smoking, alcohol intake, and BMI for all models.

### Ethical considerations

This secondary analysis used anonymised publicly available data, and was approved by the Research and Ethics committee at Instituto Nacional de Geriatría (DI-PI-005/2025).

## RESULTS

### Study population

Amongst 120,170 participants across five G2A cohorts, 24,385 participants were included in the analytic sample, contributing 78,422 longitudinal evaluations (**Supplementary Figure 1**). The weighted baseline prevalence of ADA-defined prediabetes was 50% (95%CI 48-51), ranging from 18% to 52% across cohorts. After excluding 5,340 individuals with diabetes and 474 without follow-up measurements, 18,571 participants were included in primary analyses (10,934 normoglycemic and 7,637 with prediabetes). Compared with normoglycemic participants, those with prediabetes were younger (65.02±9.97 vs. 65.86±11.01, p=0.005), predominantly female (56% vs. 53%, p<0.001), had lower education attainment (77% vs 58% below secondary; p<0.001), and exhibited a higher burden of frailty and functional deficits (frailty by Fried criteria 5.2% vs 3.8%; Frailty based on FRAIL scale 6.8% vs. 5.7%; median FI 0.16 [0.10-0.26] vs 0.13 [0.08-0.23]; all p<0.001). Having ≥1 functional limitations were also more common among participants with prediabetes at baseline compared with normoglycemic individuals, including ADL-Barthel (41% vs 32%), ADL-Katz (18% vs 16%), and IADL (17% vs 15%; all p<0.001). Median BMI was lower for individuals with prediabetes (24.8 [22.1–27.9] vs 25.3 kg/m² [22.5–28.3]; p<0.001, **Table 1**). Median follow-up at wave 1 was 3.0 years (IQR 2.9-3.7), at wave 2 7.59 years (IQR 4.3-7.8), at wave 3 11.5 years (IQR 6.1-11.9) and at wave 4 13.6 years (IQR 7.9-14.0)

**Table 1.**
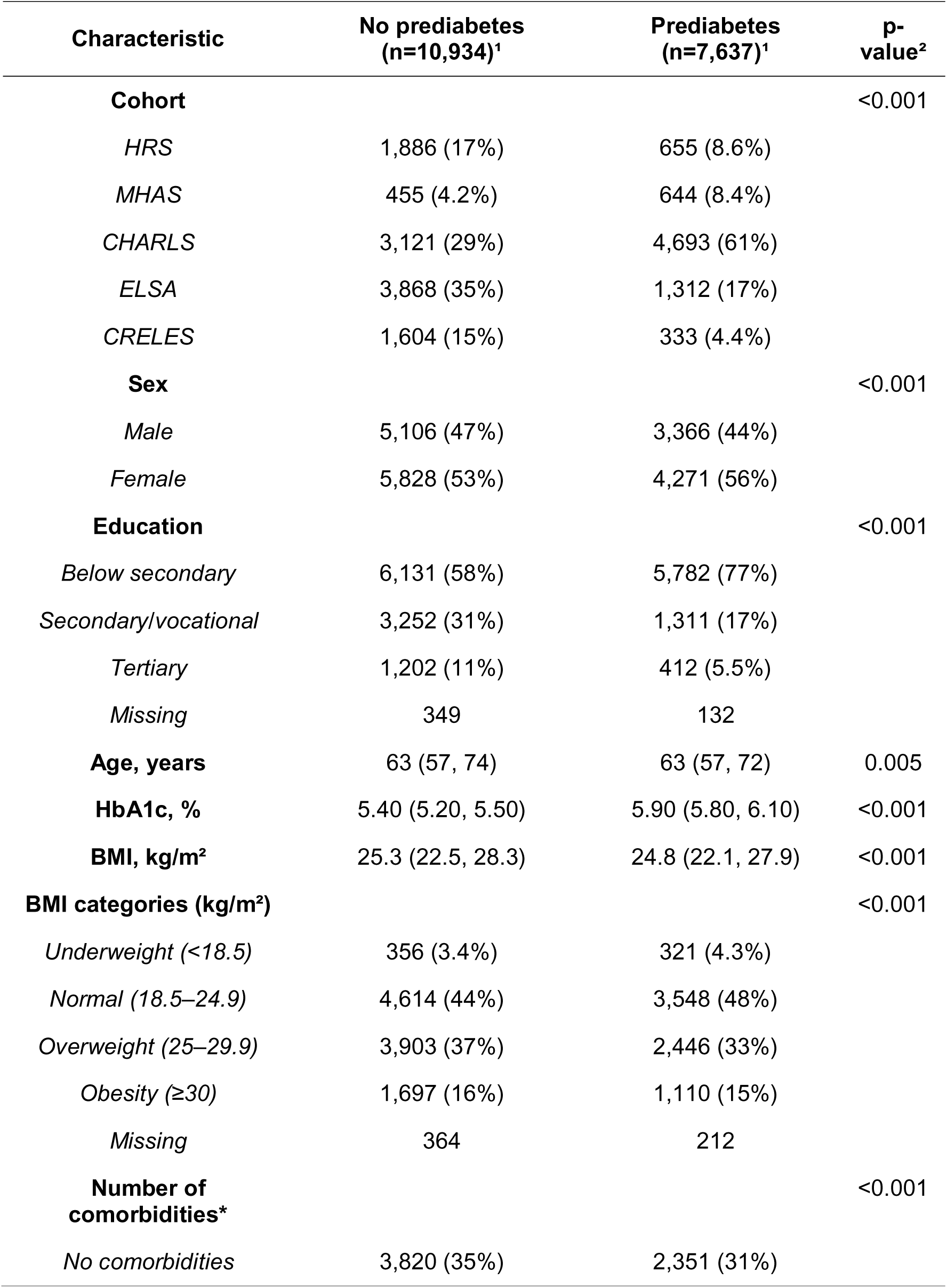
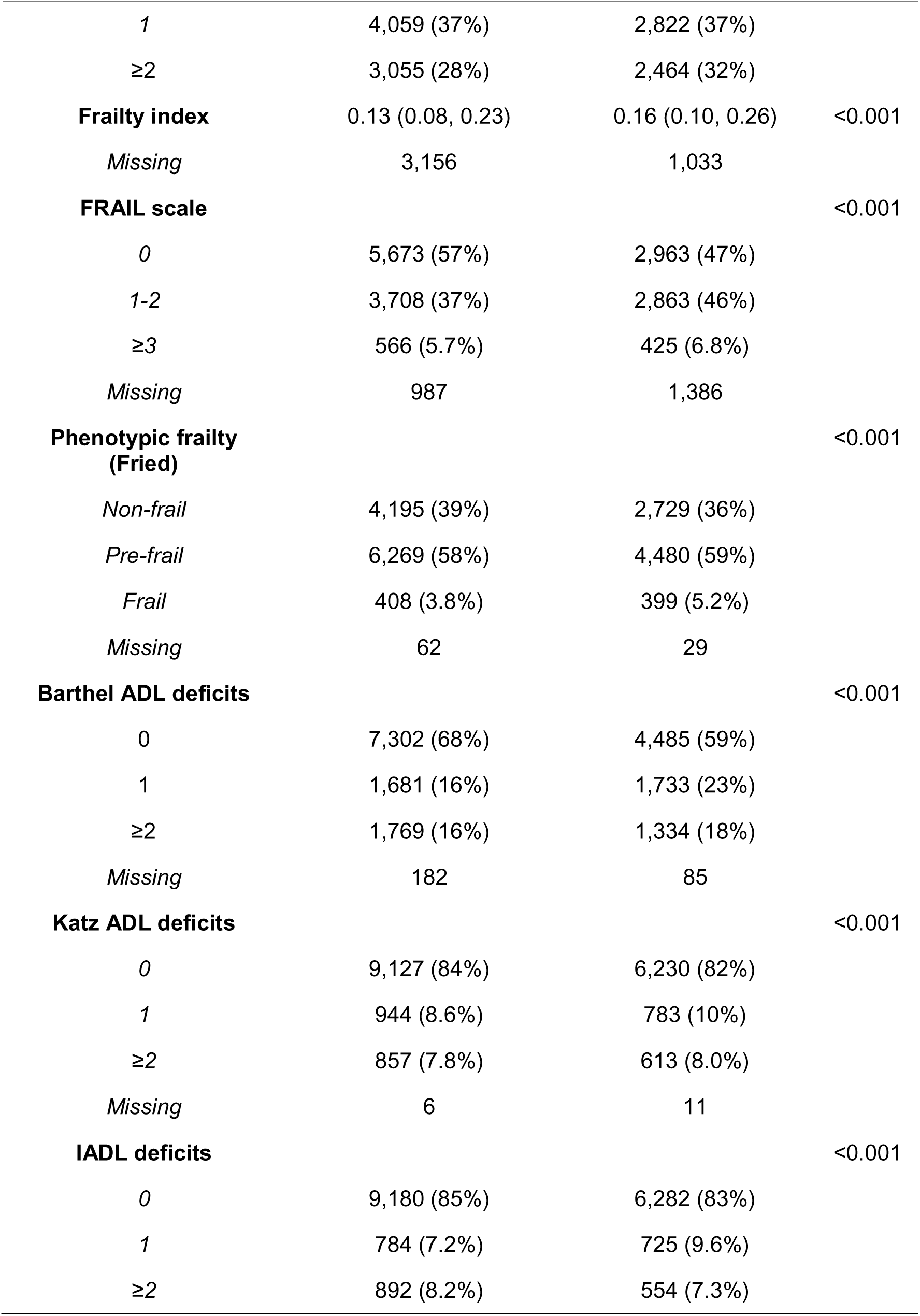
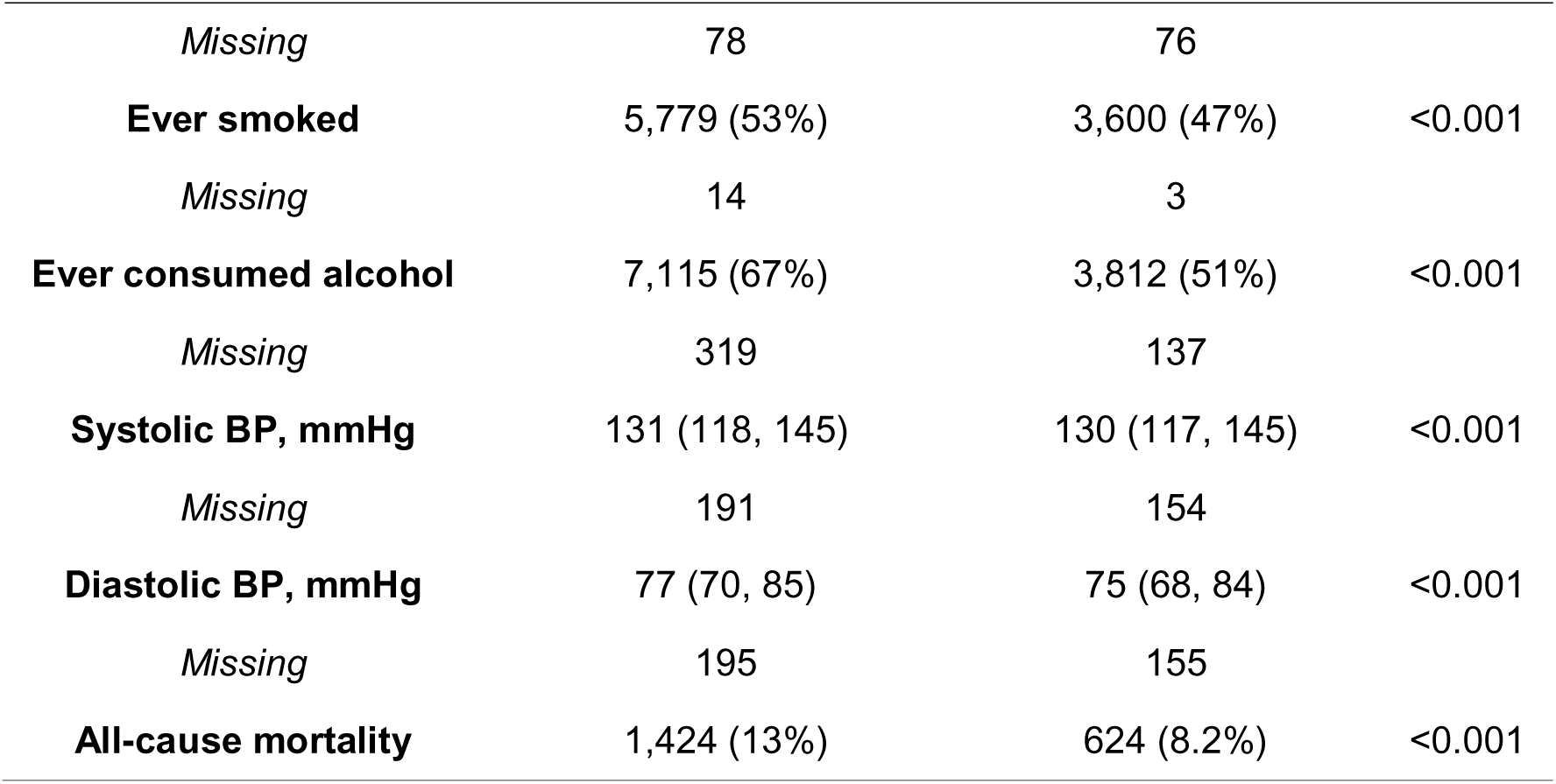
Baseline characteristics of participants with and without prediabetes with ≥1 available HbA1c measure from included G2A cohorts..

| Characteristic | No prediabetes<br>(n=10,934)<sup>1</sup> | Prediabetes<br>(n=7,637)<sup>1</sup> | p-value<sup>2</sup> |
| --- | --- | --- | --- |
| **Cohort** |  |  | <0.001 |
| *HRS* | 1,886 (17%) | 655 (8.6%) |  |
| *MHAS* | 455 (4.2%) | 644 (8.4%) |  |
| *CHARLS* | 3,121 (29%) | 4,693 (61%) |  |
| *ELSA* | 3,868 (35%) | 1,312 (17%) |  |
| *CRELES* | 1,604 (15%) | 333 (4.4%) |  |
| **Sex** |  |  | <0.001 |
| *Male* | 5,106 (47%) | 3,366 (44%) |  |
| *Female* | 5,828 (53%) | 4,271 (56%) |  |
| **Education** |  |  | <0.001 |
| *Below secondary* | 6,131 (58%) | 5,782 (77%) |  |
| *Secondary/vocational* | 3,252 (31%) | 1,311 (17%) |  |
| *Tertiary* | 1,202 (11%) | 412 (5.5%) |  |
| *Missing* | 349 | 132 |  |
| **Age, years** | 63 (57, 74) | 63 (57, 72) | 0.005 |
| **HbA1c, %** | 5.40 (5.20, 5.50) | 5.90 (5.80, 6.10) | <0.001 |
| **BMI, kg/m<sup>2</sup>** | 25.3 (22.5, 28.3) | 24.8 (22.1, 27.9) | <0.001 |
| **BMI categories (kg/m<sup>2</sup>)** |  |  | <0.001 |
| *Underweight (<18.5)* | 356 (3.4%) | 321 (4.3%) |  |
| *Normal (18.5–24.9)* | 4,614 (44%) | 3,548 (48%) |  |
| *Overweight (25–29.9)* | 3,903 (37%) | 2,446 (33%) |  |
| *Obesity ($\geq 30$)* | 1,697 (16%) | 1,110 (15%) |  |
| *Missing* | 364 | 212 |  |
| **Number of comorbidities\*** |  |  | <0.001 |
| *No comorbidities* | 3,820 (35%) | 2,351 (31%) |  |

| Characteristic | No prediabetes<br>(n=10,934)<sup>1</sup> | Prediabetes<br>(n=7,637)<sup>1</sup> | p-value<sup>2</sup> |
| --- | --- | --- | --- |
| 1 | 4,059 (37%) | 2,822 (37%) |  |
| ≥2 | 3,055 (28%) | 2,464 (32%) |  |
| **Frailty index** | 0.13 (0.08, 0.23) | 0.16 (0.10, 0.26) | <0.001 |
| *Missing* | 3,156 | 1,033 |  |
| **FRAIL scale** |  |  | <0.001 |
| 0 | 5,673 (57%) | 2,963 (47%) |  |
| 1-2 | 3,708 (37%) | 2,863 (46%) |  |
| ≥3 | 566 (5.7%) | 425 (6.8%) |  |
| *Missing* | 987 | 1,386 |  |
| **Phenotypic frailty**<br>**(Fried)** |  |  | <0.001 |
| *Non-frail* | 4,195 (39%) | 2,729 (36%) |  |
| *Pre-frail* | 6,269 (58%) | 4,480 (59%) |  |
| *Frail* | 408 (3.8%) | 399 (5.2%) |  |
| *Missing* | 62 | 29 |  |
| **Barthel ADL deficits** |  |  | <0.001 |
| 0 | 7,302 (68%) | 4,485 (59%) |  |
| 1 | 1,681 (16%) | 1,733 (23%) |  |
| ≥2 | 1,769 (16%) | 1,334 (18%) |  |
| *Missing* | 182 | 85 |  |
| **Katz ADL deficits** |  |  | <0.001 |
| 0 | 9,127 (84%) | 6,230 (82%) |  |
| 1 | 944 (8.6%) | 783 (10%) |  |
| ≥2 | 857 (7.8%) | 613 (8.0%) |  |
| *Missing* | 6 | 11 |  |
| **IADL deficits** |  |  | <0.001 |
| 0 | 9,180 (85%) | 6,282 (83%) |  |
| 1 | 784 (7.2%) | 725 (9.6%) |  |
| ≥2 | 892 (8.2%) | 554 (7.3%) |  |
| *Missing* | 78 | 76 |  |
| **Ever smoked** | 5,779 (53%) | 3,600 (47%) | <0.001 |
| *Missing* | 14 | 3 |  |
| **Ever consumed alcohol** | 7,115 (67%) | 3,812 (51%) | <0.001 |
| *Missing* | 319 | 137 |  |
| **Systolic BP, mmHg** | 131 (118, 145) | 130 (117, 145) | <0.001 |
| *Missing* | 191 | 154 |  |
| **Diastolic BP, mmHg** | 77 (70, 85) | 75 (68, 84) | <0.001 |
| *Missing* | 195 | 155 |  |
| **All-cause mortality** | 1,424 (13%) | 624 (8.2%) | <0.001 |

### Risk of functional and frailty deficits over time with a prediabetes diagnosis at baseline

Participants with prediabetes at baseline showed a higher risk of progression of deficits in ADL, IADL, FRAIL and Fried scores, and number of comorbidities compared with normoglycemic individuals (**Figure 1A**). Adjusted mixed-effects Poisson models showed a higher risk of progression for individuals with baseline prediabetes for these outcomes, with faster accumulation of deficits over time observed for ADL and IADL only (**Figure 1C**, **Table 2**). Among participants free of deficits at baseline, incident deficits in ADL, IADL, and number of comorbidities were higher from the first follow-up wave onward in individuals with prediabetes compared with normoglycemia without significant time by prediabetes interactions, indicating incident deficit rates did not differ in their trajectory over time (**Figure 1B-C**, **Supplementary Table 5**). Analyses using fixed-effects Poisson models across follow-up waves yielded similar findings, even after including additional adjustment for smoking, alcohol intake, and BMI (**Supplementary Figure 2**).

**Figure 1.**
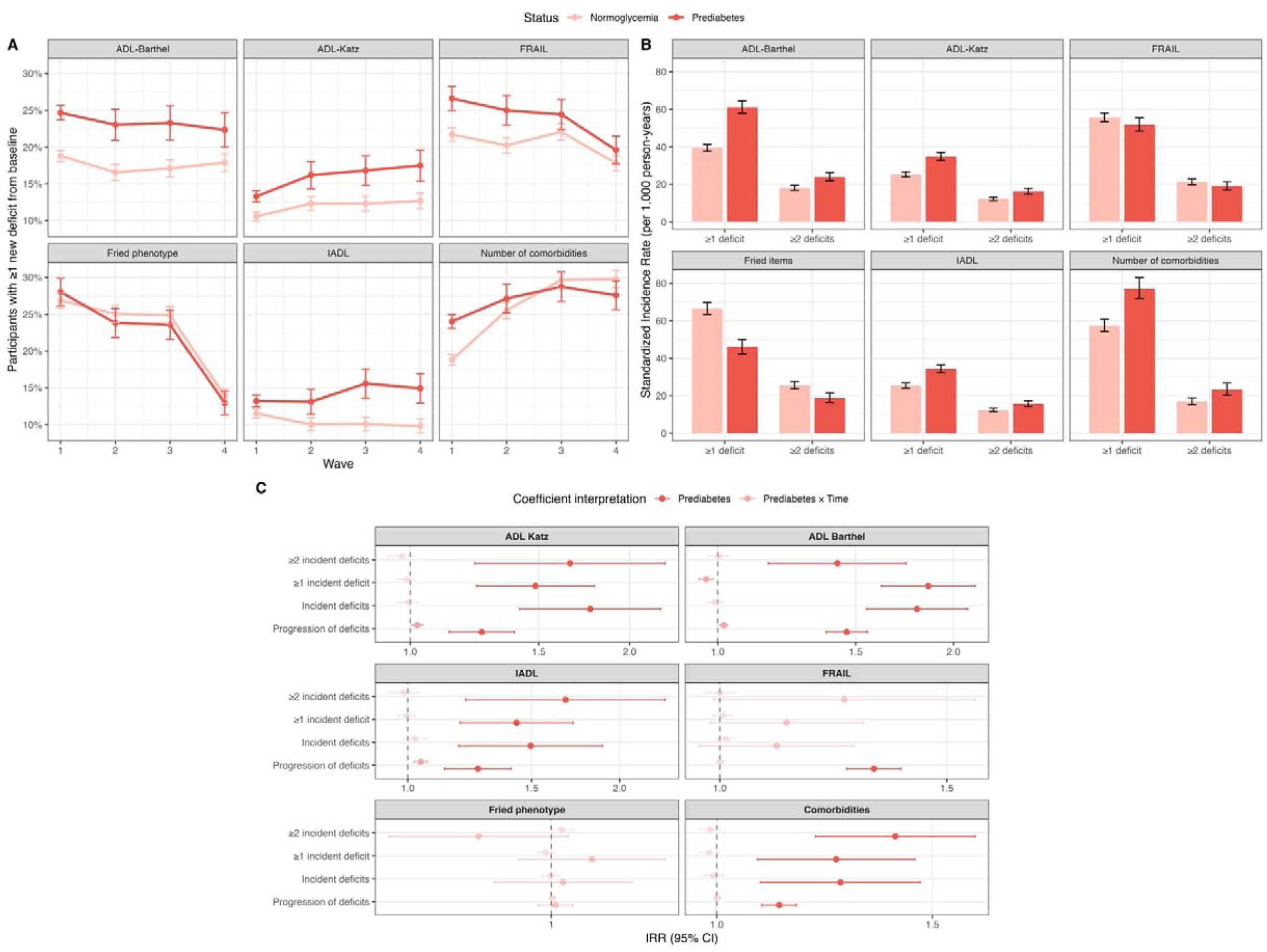
**(A)** Weighted prevalence of participants with ≥1 new deficit from baseline for all evaluated outcomes amongst participants without diabetes, stratified by prediabetes and normoglycemia. (**B**) Among participants without deficits at baseline, age- and sex-standardized incidence of ≥1 and ≥2 deficits for all evaluated outcomes, stratified by prediabetes and normoglycemia. (**C**) Mixed effects Poisson regression models with robust standard errors assessing the risk of progression and incidence of deficits in functional outcomes and frailty amongst individuals with prediabetes and a prediabetes by time interaction compared to normoglycemia. All analyses included participants without diabetes from HRS, ELSA, CRELES, MHAS and CHARLS.

**Table 2.**
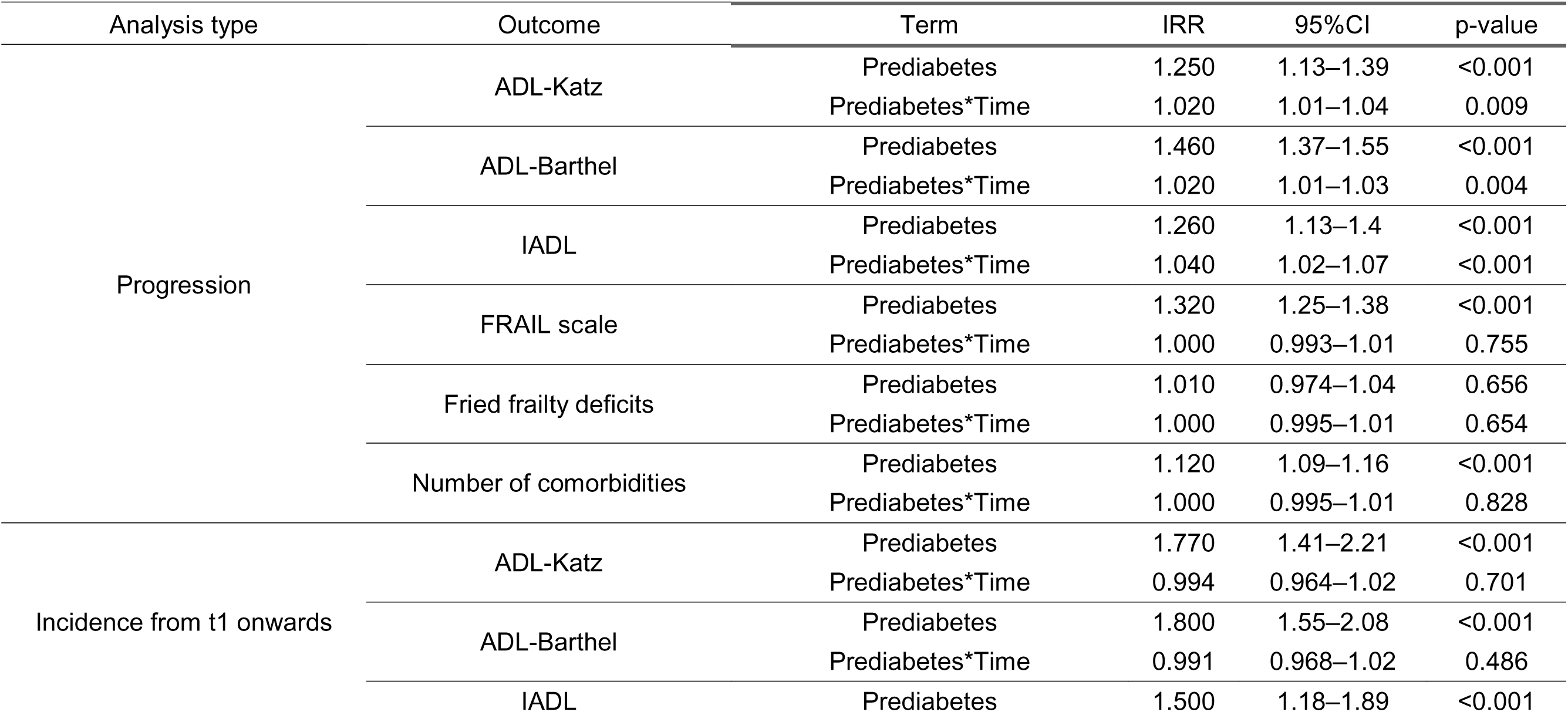
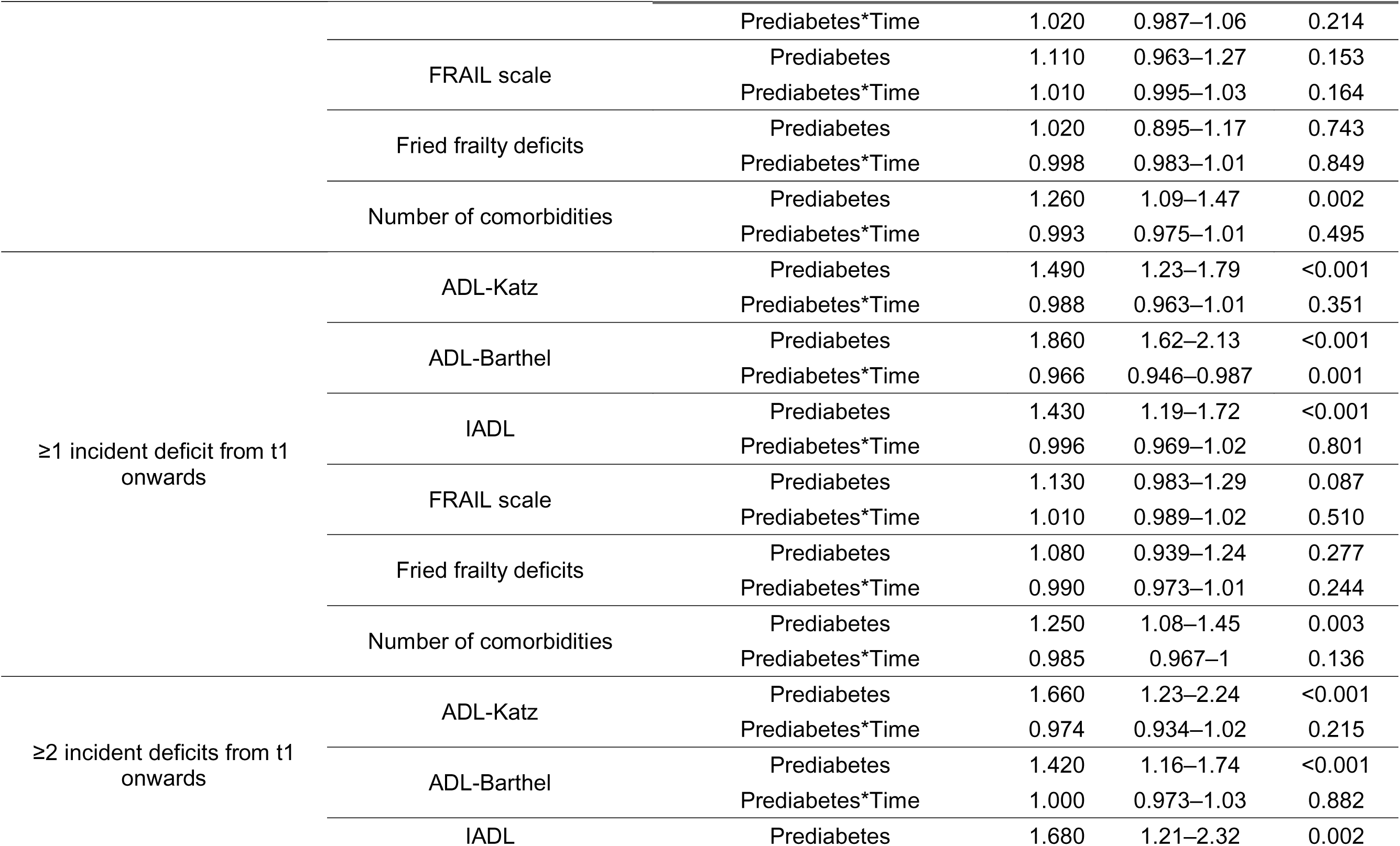
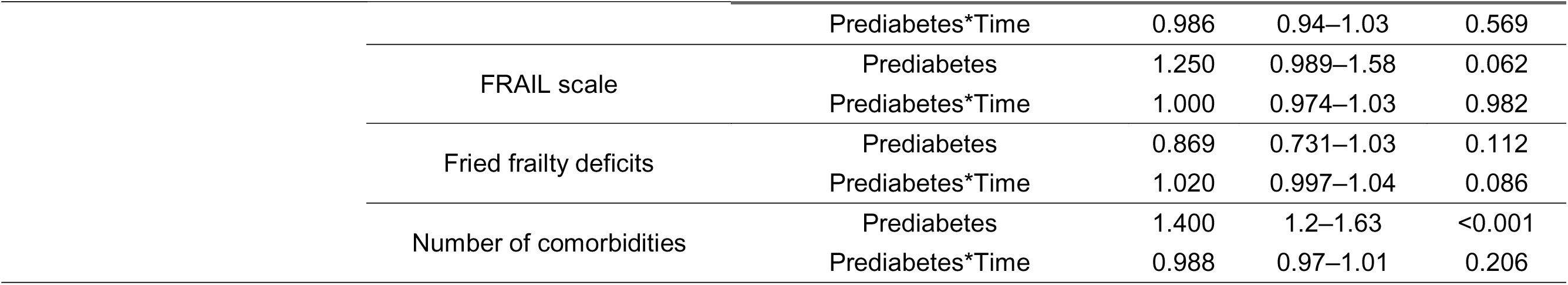
Mixed effects Poisson regression models assessing the risk of progression and incidence of deficits in ADL, IADL, FRAIL, Fried frailty phenotype and number of comorbidities associated to a single diagnosis of prediabetes at baseline compared to normoglycemic individuals amongst participants without diabetes from included G2A cohorts. ***<u>Abbreviations:</u>*** *ADL, Activities of daily living; IADL, instrumental activities of daily living; FRAIL scale, F - Fatigue R - Resistance A - Ambulation I - Illnesses L - Loss of weight; IRR, Incidence rate ratio; 95%CI, 95% confidence interval* ***Note:*** *For progression models we assessed participants with deficits at baseline and the prediabetes coefficient is interpreted as differences at baseline; in contrast, for incidence models we assessed participants without deficits in each respective category, and the prediabetes coefficient is interpreted as incidence starting at the first follow-up wave*.

### Frailty index progression over time with a prediabetes diagnosis at baseline

Over time, participants with a single diagnosis of prediabetes at baseline consistently exhibited higher FI values than normoglycemic individuals. In models adjusted for age, sex, and cohort of origin, prediabetes was associated with higher FI values across all waves (β=0.020, 95%CI 0.016-0.024). A prediabetes by time interaction indicated an increased rate of FI progression compared with normoglycemic participants after five years of follow-up (β=0.001, 95%CI 0.001-0.002, **Supplementary Figure 3, Supplementary Table 5**).

### Glycemic transitions over time

Glycemic transitions were evaluated among participants with ≥2 HbA1c measurements over time from ELSA, HRS, and CRELES (n=7,840 contributing 21,149 longitudinal evaluations, **Supplementary Figure 1**). Normoglycemic participants decreased from 64.1% of participants at baseline to 41.1% at their last measured HbA1c, prediabetes increased from 21.3% to 29.8%, and diabetes from 14.6% to 29.1% at follow-up (**Figure 2A**). When accounting for death over the full follow-up, amongst those with normoglycemia at baseline 11.7% progressed to diabetes, 27.5% progressed to prediabetes, and 10.4% died; similarly, for those with prediabetes 25.5% progressed to diabetes, 20.9% regressed to normoglycemia, and 13.1% died. Amongst those with diabetes 20.8% died over the follow-up (**Figure 2B**). Among those with wave 1 with available data at wave 2 (n=7,840) the proportion of normoglycemic individuals decreased from 64.1% to 37.4%, while prediabetes and diabetes increased to 35.5% and 27.1%, respectively. Among participants with available measures between waves 2-3 (n=4,083), normoglycemia increased from 45.1% to 48.3%, prediabetes declined from 39.3% to 32.8%, and diabetes increased from 15.6% to 18.9%. Finally, amongst participants with data between waves 3-4 (n=1,386), normoglycemia rose from 50.9% to 62.2%, prediabetes decreased from 37.7% to 24.4%, and diabetes increased from 11,5% to 13.4% (**Figure 2C-E**). Among participants with prediabetes, progression to diabetes decreased across successive waves from 24.3% to 4.6%, whereas regression to normoglycemia increased from 20.8% to 44.1%. Similarly, among normoglycemic participants, progression to prediabetes declined from 37.2% to 9.9%, and progression to diabetes from 11.3% to 0.4% (**Supplementary Table 6**).

**Figure 2.**
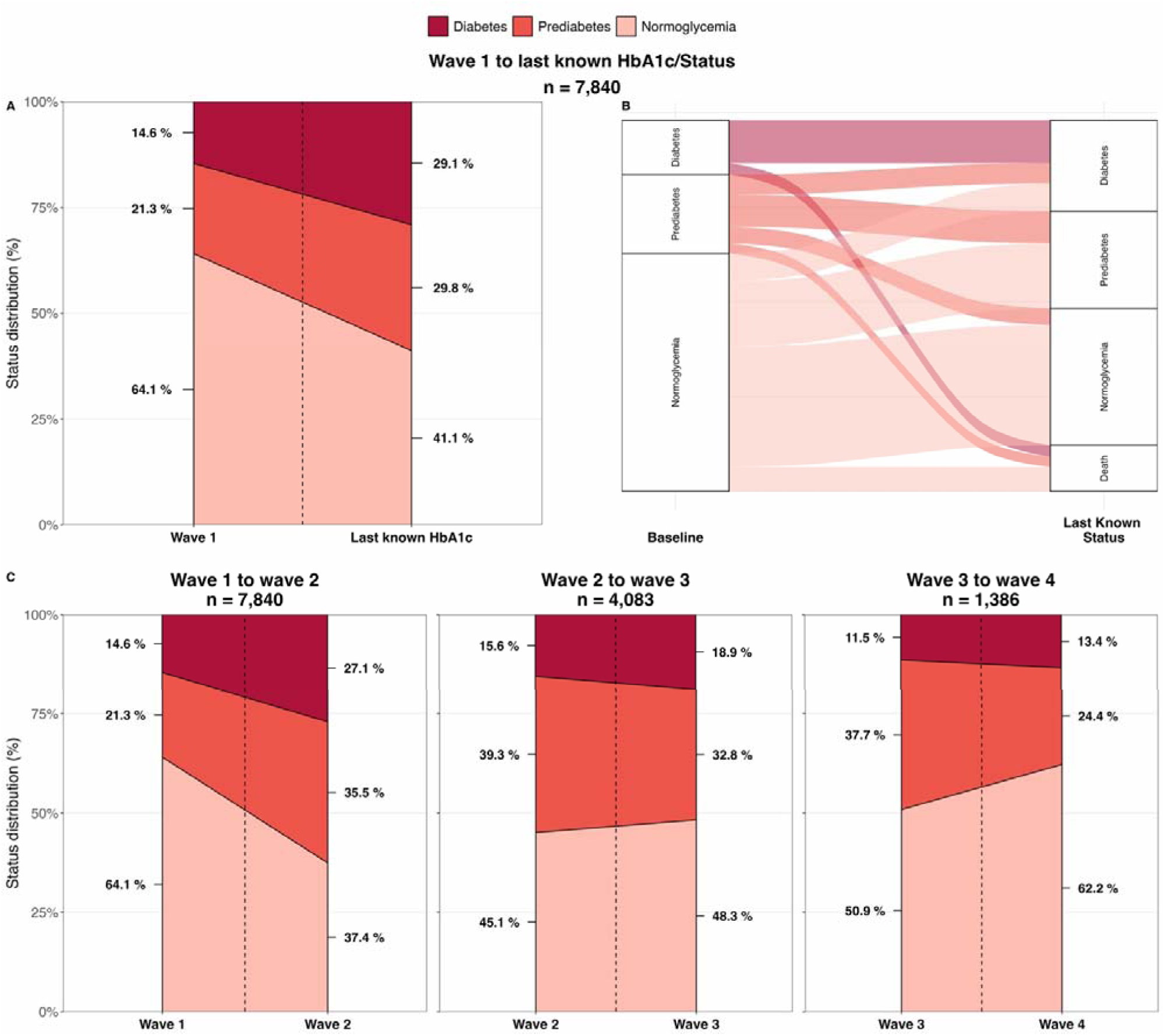
Transitions in glycemic status across longitudinal assessments among participants with ≥2 HbA1c measures from CRELES, HRS, and ELSA, showing transitions from normoglycemia, prediabetes, and diabetes to their last known glycemic status over the full follow-up time (**A**), and after accounting for competing risk of death (**B**). The figure also compares participants from the prior to the next wave with available HbA1c measures to assess shorter term transitions (**C**). Diabetes was treated as a non-reversible state for all analyses.

### Glycemic status and risk of functional deficits over time

Among participants without baseline deficits, participants with diabetes exhibited the highest rates of incident deficits across all outcomes, whereas individuals with prediabetes generally showed higher risk compared with normoglycemic participants, particularly during earlier follow-up waves (**Figure 3A**). GEE models confirmed that both prediabetes and diabetes were longitudinal predictors of incident deficits across all evaluated outcomes and, although absolute risk remained elevated, the relative differences in incidence rates between glycemic groups decreased slightly over the follow-up period (**Figure 3B**, **Table 3**). Regarding progression of existing deficits, only diabetes was associated with significantly higher rates of worsening IADL and FRAIL deficits, as well as increasing number of comorbidities. Prediabetes was only associated with higher risk of comorbidity accumulation (**Figure 3B**, **Table 3**). In a subanalysis restricted to individuals with prediabetes at baseline (n=1,670), regression to normoglycemia was associated with increased risk of incident deficits in ADL, FRAIL and Fried. Progression to diabetes was associated with decreased risk of ADL and Fried deficits, without changes in risk over time (**Supplementary Table 9**). Findings remained largely similar when performing additional adjustments for smoking, alcohol intake and time-varying BMI, except for ADL (**Supplementary Table 10**).

**Figure 3.**
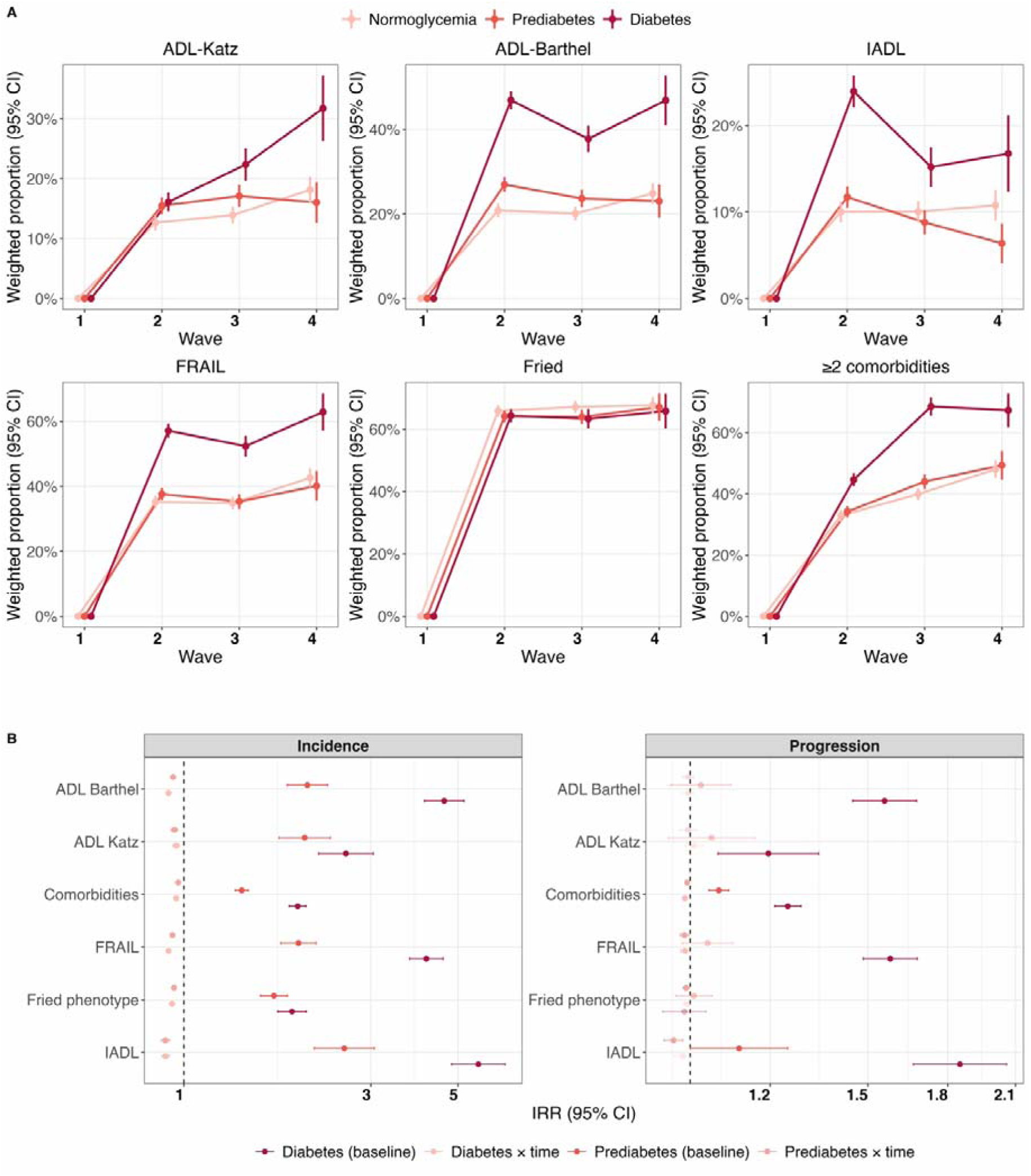
**(A)** Weighted cumulative incidence of ≥1 deficit in ADL-Katz, ADL-Barthel, IADL, FRAIL and Fried, or ≥2 comorbidities over successive study waves among participants with ≥1 HbA1c measure over time from CRELES, HRS, and ELSA. **(B)** Generalized estimating equation models with a Poisson link assessing risk of incidence and progression in deficits for individuals with diabetes and prediabetes overall and with a time interaction compared to individuals with normoglycemia among participants with ≥1 HbA1c measure over time from CRELES, HRS, and ELSA.

**Table 3.**
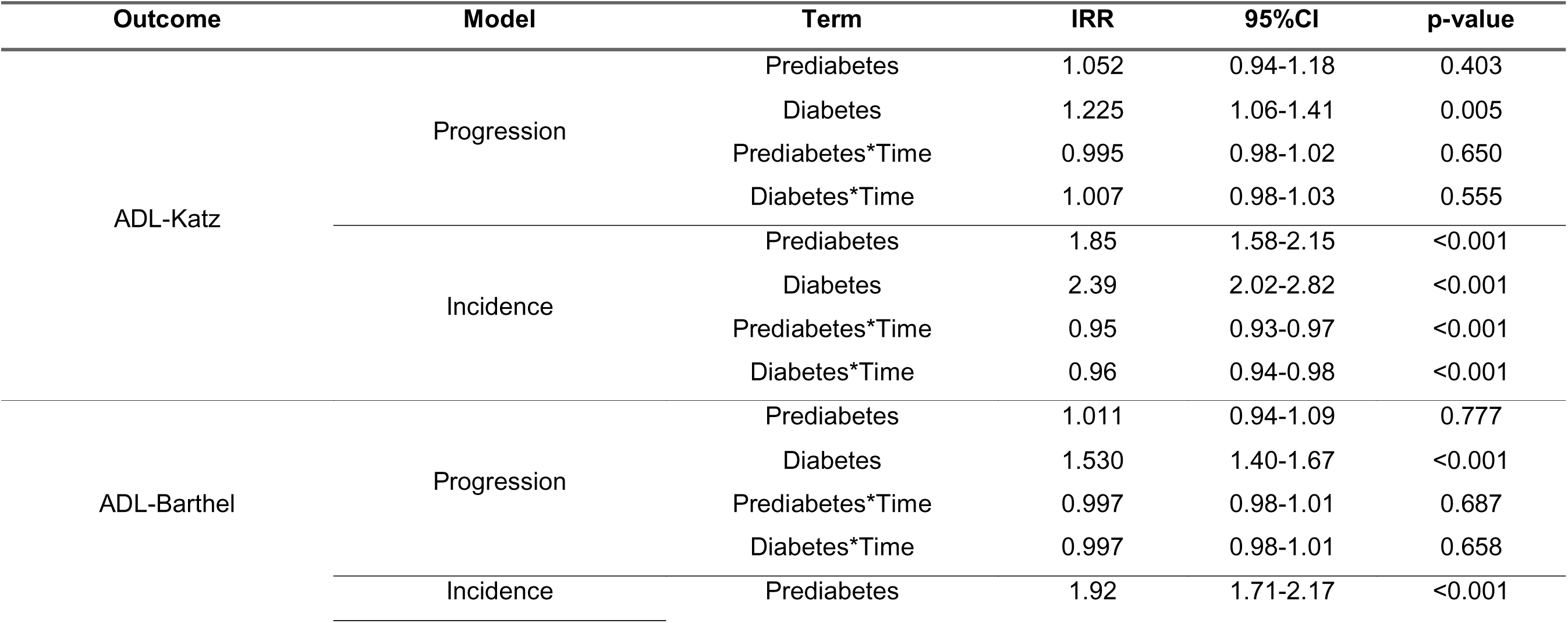
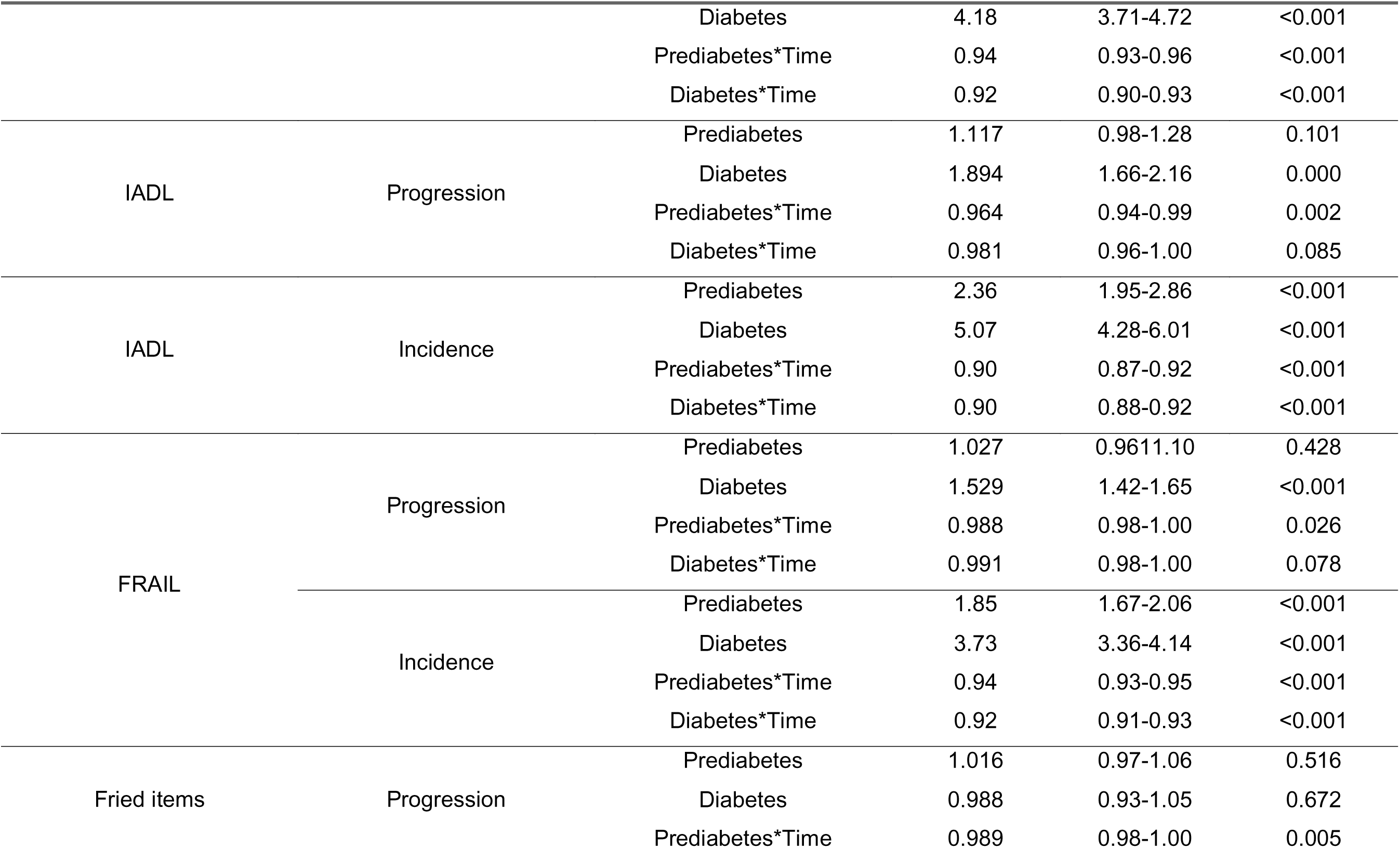
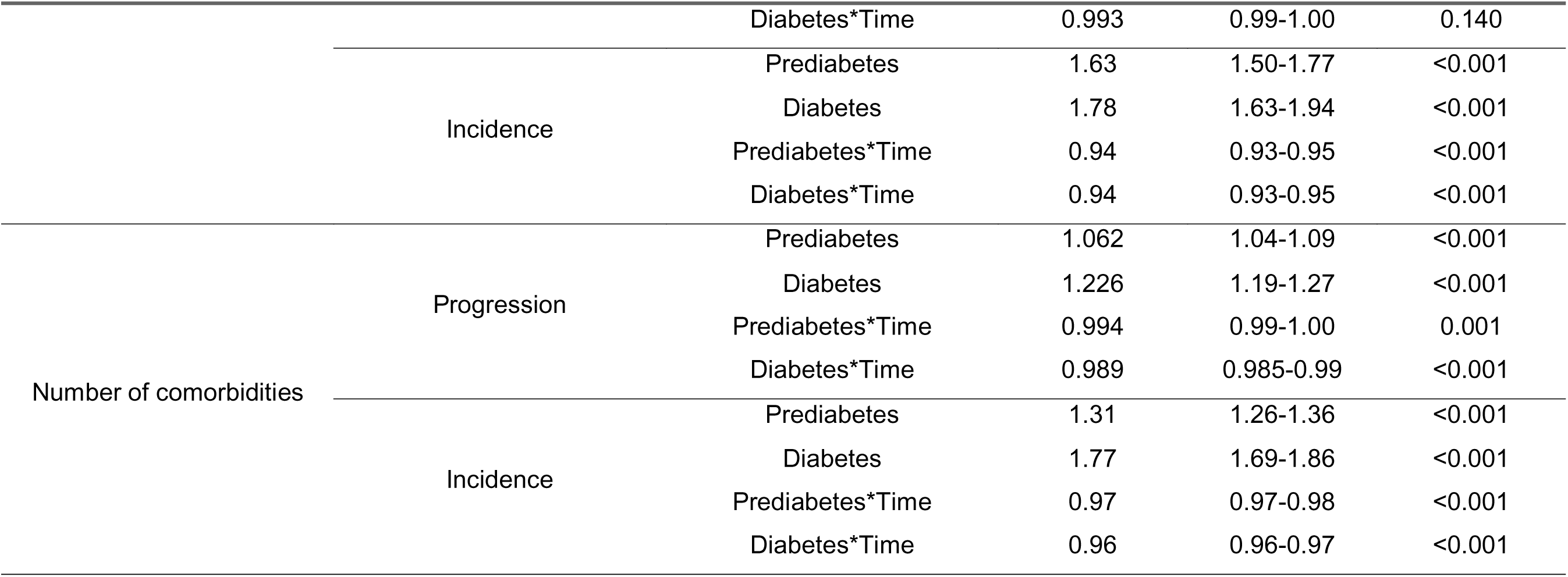
Generalized Estimating Equation models with a Poisson link assessing the risk of progression and incidence of deficits in ADL, IADL, FRAIL, Fried frailty and number of comorbidities associated to diagnosis of prediabetes or diabetes at all evaluated time points compared to normoglycemia amongst participants from ELSA, HRS and CRELES with ≥1 HbA1c measurement over time. ***<u>Abbreviations:</u>*** *ADL, Activities of daily living; IADL, instrumental activities of daily living; FRAIL scale, F - Fatigue R - Resistance A - Ambulation I - Illnesses L - Loss of weight; IRR, Incidence rate ratio; 95%CI, 95% confidence interval* ***Note:*** *For progression models we assessed participants with deficits at baseline, and for incidence models we assessed participants without deficits in each respective category. Models were adjusted for age, cohort and sex*.

### Glycemic transitions and frailty index progression

Individuals with diabetes exhibited the highest burden of deficit accumulation and the steepest rate of FI progression over time (**Figure 4A**). Compared with normoglycemic participants, diabetes was associated with higher FI values (β=0.025, 95% CI 0.018-0.032) and a significant interaction with time indicating an accelerating trajectory. Prediabetes was associated with a smaller increase in FI (β=0.005, 95% CI 0.001-0.010) and a negative interaction with time. Transition analyses further showed that, among participants with prediabetes, progression to diabetes was associated with a rapid increase in FI (β=0.033, 95% CI 0.018-0.049), while regression to normoglycemia was associated with higher FI levels compared to stable prediabetes (β=0.023, 95% CI 0.001-0.045), with stable long-term trajectories (**Figure 4B**).

**Figure 4.**
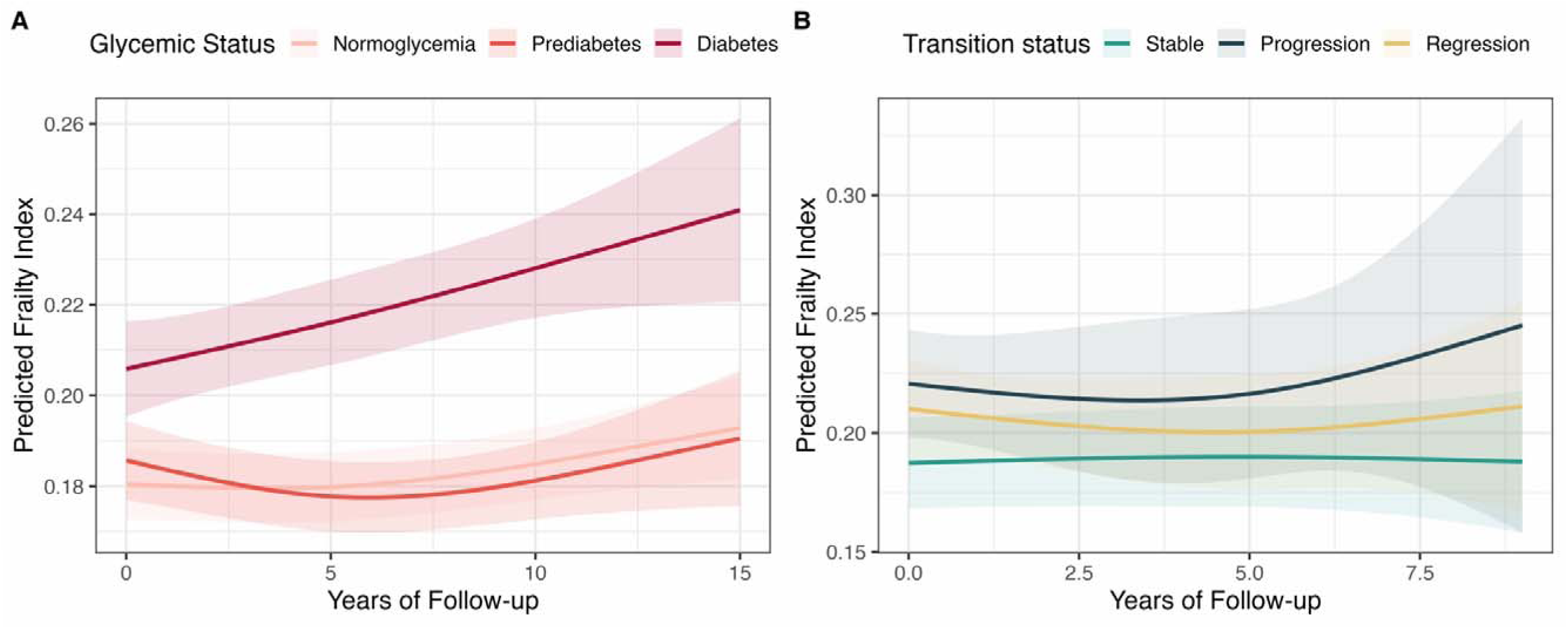
Predicted frailty index trajectories over time derived from generalized estimating equation models with a Gaussian link stratified by glycemic status among participants with ≥1 HbA1c measure over time from CRELES, HRS, and ELSA (**A**), and stratified by glycemic transitions among participants with prediabetes and ≥1 HbA1c measure over time from CRELES, HRS, and ELSA (**B**).

## DISCUSSION

In this secondary analysis of harmonized longitudinal data from five international cohorts involving adults aged ≥50 years, we characterized prediabetes as a risk factor for functional decline, frailty, and deficit accumulation across multiple measures of physical function. These associations were observed both when prediabetes was defined at a single time point and when modeled dynamically over time while accounting for the risk of progression to diabetes and regression to normoglycemia. While diabetes conferred the greatest risk across all outcomes, prediabetes was independently associated with elevated risk compared with normoglycemia. Notably, among individuals with prediabetes, regression to normoglycemia was associated with higher rates of functional decline and frailty, whereas progression to diabetes was not associated with a corresponding increase in risk relative to stable prediabetes. This is striking, as regression to normoglycemia was common in our cohort likely due underlying unintentional weight loss or sarcopenia, which may mediate the observed associations. Our findings suggest that prediabetes in older adults may reflect a broader adaptive metabolic state related to physiological vulnerability, rather than solely an intermediate stage in the progression to diabetes. Age-related alterations including insulin resistance, mitochondrial dysfunction, chronic low-grade inflammation, and loss of skeletal muscle mass are likely to contribute both to dysglycemia and to declining physiological reserve^18,19^. Based on our findings, we propose that prediabetes in older adults may be useful as a clinical marker of reduced resilience, identifying individuals at increased risk of frailty and functional impairment independent of its role as a risk factor for progression to overt diabetes.

Our findings extend previous evidence linking prediabetes to adverse health outcomes by demonstrating consistent associations between prediabetes and incresaed risk of deficit accumulation across multiple domains of functional status and frailty using multinational harmonized longitudinal data. Prior studies have reported heterogeneous associations that vary by age, population, and outcome definition. For example, prediabetes has been associated with increased risk of disability and mortality in middle-aged Japanese adults over long-term follow-up^20^, whereas among older Chinese participants it has been associated to prevalent but not incident frailty^21^. Similarly, studies from the US and Mexico suggest that prediabetes confers increased mortality in midlife but not in older age^22,23^, while evidence from adults ≥80 years in Israel indicates lower all-cause mortality but higher cardiovascular risk among those with prediabetes compared to normoglycemia^24^. This heterogeneity may be partly attributable to limitations of studies relying on a single assessment of glycemic status, particularly in older adults, in whom glycemic trajectories are shaped by competing risks, selective survival, and underlying health status. Moreover, exposure time to dysglycemia is difficult to assess even in longitudinal settings, and its impact on long-term metabolic and functional outcomes has been insufficiently characterized in older adults^25^. By incorporating both baseline and time-varying glycemic measures, our study captures the dynamic nature of dysglycemia in older adults. In addition, the use of multiple complementary frailty constructs, including phenotype-based and deficit-accumulation approaches, enables a more comprehensive characterization of aging-related vulnerability. Our findings support the conceptualization of prediabetes as part of a broader metabolic aging phenotype rather than solely an intermediate stage in the development of diabetes, and contribute to the understanding of its associated risk of physical disability, multimorbidity, and frailty^26^.

Evidence from studies incorporating dynamic measures of glycemic status further supports the importance of considering glycemic trajectories rather than single timepoint classifications when assessing risk associated with prediabetes. A prior analysis of ELSA using repeated measures of glycaemia demonstrated that higher HbA1c levels and diabetes are associated with steeper trajectories of frailty progression, which may exemplify the cumulative impact of chronic dysglycemia on physiological reserve^27^. Results from the Swedish National Study on Aging and Care in Kungsholmen showed that prediabetes was associated with an increased chair stand time, decreased walking speed and accelerated disability progression, even when accounting for progression to diabetes^10^. Among adults with prediabetes, progression to diabetes has also been associated with clinically meaningful declines in strength, mobility, and balance over time^28^. Notably, the relationship between prediabetes with physical function and frailty appers to be bidirectional. Frailty itself appears to influence glycemic trajectories, with frail individuals with prediabetes showing a higher risk of progression to diabetes and markedly increased risks of cardiovascular events and mortality compared with non-frail individuals^29^. Conversely, in the oldest-old, frailty has been associated with a lower risk of progression to diabetes and a higher probability of remaining stable or regressing to normoglycemia, consistent with patterns of metabolic decline and selective survival^24^. These findings support a model in which frailty and prediabetes are interconnected, with underlying processes such as sarcopenia, inflammation, and multimorbidity shaping both metabolic trajectories and functional outcomes. This evidence aligns with our findings, emphasizing a need to move beyond static definitions of prediabetes and instead consider glycemic status as a dynamic process embedded within the broader context of aging biology.

Our observation that regression to normoglycemia amongst older adults with prediabetes was associated with increased risk of frailty is counterintuitive and warrants cautious interpretation. We hypothesize that rather than indicating a detrimental effect of glycemic improvement, this finding likely reflects underlying processes associated with declining health status. In older adults, reductions in HbA1c may occur in the context of unintentional weight loss, sarcopenia, malnutrition, or chronic disease, all of which are established contributors to frailty and disability^30–32^. Thus, regression to normoglycemia and more broadly HbA1c variability may represent a marker of physiological deterioration and increased frailty risk rather than metabolic recovery in older adults with prediabetes^33^. Although our analyses adjusted for time-varying BMI and lifestyle factors to reduce the likelihood of weightloss and age-related loss of muscle mass influencing the observed associations^34^, residual confounding and reverse causation cannot be excluded. Further studies incorporating longitudinal measures of body composition, muscle mass and function, nutritional status, and inflammatory markers are needed to better characterize the mechanisms underlying the impact of glycemic transitions on functional outcomes and frailty risk.

Our study has several strengths, including the use of a large nationally representative sample of older adults, extended longitudinal follow-up, and inclusion of multinational aging studies contribute to the external validity of our findings across different geographical, ethnic, and socioeconomic contexts. Data harmonization across G2A studies enabled broadly consistent definitions of exposures and outcomes, and the use of longitudinal modelling approaches accounted for within-individual changes over time. Moreover, we were able to assess two complementary approaches to assess the impact of prediabetes and glycemic status on frailty and functional outcomes, including a single diagnosis of prediabetes, and glycemic transitions over time. However, we acknowledge some limitations which should be considered to adequately interpret our results. First, although G2A studies can be pooled and harmonized for most variables and outcomes, we were unable to construct a FI with the same number of items across all cohorts; specifically, although we constructed a 31-item FI for MHAS, HRS, ELSA and CHARLS, some unavailable items in CRELES only allowed us to construct a 26-item FI for this cohort. To address this, we used mixed effects models and adjusted for study cohort to handle between-center heterogeneity, and additionally conducted FI analyses excluding CRELES, observing overall similar results. Second, due to the observational design of all G2A studies, unmeasured factors such as unintentional weight loss, sarcopenia, or underlying illness may influence the observed associations, especially for glycemic transitions over time. We partially addressed this by performing adjustment by time-varying BMI; however, no objective measures of muscle mass were available across all cohorts. Third, glycemic status and prediabetes were assessed using only HbA1c as glucose or oral glucose tolerance test data were not available, which may affect adequate assessment of glycemic status; notably, HbA1c values may be unreliable in older adults as they may be affected by age-related changes in erythrocyte turnover and comorbid conditions, affecting their reliability to assess glycemic status over time^35^. Finally, analyses of glycemic transitions were restricted to a subset of cohorts with repeated HbA1c measurements, which may introduce selection and survivorship bias, leading only overall healthier participants to be included in this cohort and potentially influence reliability of results and limit comparability.

## Conclusion

In this large multinational longitudinal analysis of older adults, prediabetes was associated with accelerated functional decline, increased frailty burden, and greater deficit accumulation across multiple domains. These associations were evident both when prediabetes was defined at a single time point and when glycemic status was modelled dynamically and persisted independently of progression to overt diabetes and adjustment for time-varying BMI. Notably, regression to normoglycemia among individuals with prediabetes was associated with a higher risk of functional deterioration, suggesting that improvements in glycemic markers in later life may reflect underlying physiological decline rather than true metabolic recovery. In contrast, progression to diabetes did not uniformly confer additional risk beyond stable prediabetes for several functional outcomes. Our findings challenge th conventional paradigm that frames prediabetes primarily as a precursor to diabetes. Instead, they support its interpretation in older adults as a marker of systemic metabolic vulnerability and reduced physiological reserve, especially when assessed using HbA1c. Incorporating glycemic status into geriatric risk stratification may improve identification of individuals at high risk of frailty and functional decline and may be complementary to assessment of nutritional status. Further research should characterize mechanisms linking dysglycemia with aging and clarify the clinical implications of glycemic transitions in older life.

## Data sharing statement

All code and materials regarding implemented G2A studies are available for reproducibility of results http://github.com/oyaxbell/prediabetes_g2a/.

## Author contributions

Andrea Malagon-Liceaga: Methodology, formal analysis, visualization, writing - original draft, writing-review and editing. Martín Roberto Basile-Alvarez: Methodology, software, visualization, formal analysis. Carlos A. Fermín-Martínez: Methodology, formal analysis, data curation, visualization, writing - review and editing. Diana Laura Ramírez-Rivera: Data curation, investigation, writing - review and editing. Jerónimo Perezalonso-Espinosa: Data curation, investigation, writing - review and editing. Juan Pablo Díaz-Sánchez: Data curation, investigation, writing - review and editing. Sara García-González: Investigation, writing - review and editing. Karime Berenice Carrillo-Herrera: Investigation, writing - review and editing. Leslie Alitzel Cabrera Quintana: Investigation, writing-review and editing. Neftali Eduardo Antonio-Villa: Methodology, writing-review and editing. Natalia Gomes Gonçalves: Methodology, writing-review and editing. Carmen Garcia-Peña: Methodology, supervision, conceptualization, writing-review and editing. Omar Yaxmehen Bello-Chavolla: Conceptualization, methodology, supervision, writing-original draft, writing-review and editing. All authors contributed important intellectual content during manuscript drafting or revision, had access to the data, and accepted accountability for the overall work by ensuring that questions related to the accuracy or integrity of any part of the work were appropriately investigated and resolved.

## Data Availability

https://g2aging.org/home

## Acknowledgments

Andrea Malagon-Liceaga was supported by scholarship from the Secretaría de Ciencia, Humanidades, Tecnología e Innovación (SECHITI scholarship number 2032190). This project was registered and approved by the Research Committee at Instituto Nacional de Geriatría, project number DI-PI-005/2025. CAFM, JPDS, and JPE are enrolled at the PECEM Program of the Faculty of Medicine at UNAM and are supported by SECIHTI. MBA is also supported by SECIHTI.

