## Supplementary Material for "Prediabetes and glycemic transitions as determinants of frailty and functional decline in adults aged 50 years and older: A longitudinal analysis from five multinational aging cohorts"

### SUPPLEMENTARY METHODS

*Sampling methods for all included G2A studies*

**MHAS:** The initial 2001 sample included individuals born before 1951, selected from the National Employment Survey (ENE), representing rural and urban areas in the 32 federal entities, with oversampling in six states with high migration to the United States. In 2012, a new cohort of people born between 1952 and 1962 was incorporated, along with their spouses or partners, selected from the National Survey of Occupation and Employment (ENOE). In 2018, another cohort born between 1962 and 1968 was added. Additionally, in 2012, a random subsample was selected for the collection of biomarkers, anthropometric measurements, and physical performance tests.

**CRELES:** The original cohort (pre-1945) employed a national sampling strategy designed to represent Costa Rican residents aged 60 years or older in 2005, regardless of nationality. The selection process began with a random sample of 9,600 individuals from the 2000 Census, stratified into five-year age groups to ensure representation. A subsample of 60 out of the 102 health areas in the country was selected, covering 59% of the national territory. For the longitudinal study, approximately 5,000 people were identified, of whom 2,827 were finally interviewed. Response rates were affected by factors such as mortality (19%) and difficulties in locating participants (18%). The retirement cohort (1945-1955) was not included in our study because it contains different participants from those in the original CRELES cohort.

**ELSA:** Sampling strategy for adults aged 50 years or older living in private households in England. The initial sample was drawn from participants of the Health Survey for England (HSE) between 1998 and 2001, selecting those who met the age criterion and had given consent, stratified into five-year age groups. This initial sample included 11,578 households and 18,813 individuals. In waves 3, 4, 6, 7, 9, and 10, "refreshment" samples were incorporated from other years of the HSE, obtained by selecting households where at least one member met the age criteria (50 years or older). Each refreshment sample is incorporated into the general cohort and included in subsequent waves. Additionally, anthropometric information is available in waves 2, 4, and 6 (2004, 2008, and 2012). In waves 8 and 9, anthropometric data were collected for 50% of the participants in each wave. Within the framework of ELSA, data collection is structured in sequential biennial waves, alternating between core waves of standardized interviews and complementary waves with biomedical assessments by nursing staff. Specifically, waves 3 (2006-2007), 5 (2010-2011), and 7 (2014-2015) constitute core interview rounds without biomedical components, resulting in the absence of anthropometric measurements (height, body weight, BMI, gait speed, and grip strength) fundamental for constructing frailty and functionality indices. These biomedical data are obtained exclusively in waves with nursing visits, given that they involve invasive or semi-invasive procedures such as direct anthropometry and biological sample extraction for glycemic analysis. Including odd-numbered waves would generate incomplete datasets for all participants. Therefore, exclusion was chosen to preserve longitudinal integrity, minimize imputations, mitigate information bias, and ensure the completeness of covariates.

**CHARLS:** It uses a four-stage sampling design, representative of adults aged 45 years or older residing in 28 provinces. In the first stage, 150 counties were selected through random stratified sampling by region, level of urbanization, and GDP per capita. In each county, three primary sampling units (PSUs) were chosen with probability proportional to size (PPS). Subsequently, dwellings were listed using GIS software and randomly selected 80 households per PSU, adjusting the proportion according to age eligibility. Within each household, individuals aged 45 years or older were randomly selected, along with individuals aged 40–44 years as a refreshment sample for future waves. In 2012, a pilot study was conducted to evaluate and adjust the sampling methodology, incorporating 2,385 individuals from 1,554 households in Zhejiang and Gansu. For this study, subjects under 50 years of age were filtered.

**HRS:** It employs a probabilistic multi-stage area-based design, representative of U.S. adults aged 51 to 61 years (born between 1931 and 1941) residing in private households. It includes oversamples of Black individuals, Hispanics, and Florida residents. Selection was conducted at the level of Metropolitan Statistical Areas or counties (primary stage), with geographic stratification and specific weights for oversampled subpopulations. The design incorporates post-stratification to adjust demographic distributions to the 1990 Census. Anthropometric information is collected in waves 8 to 14, alternating between two halves of the sample (one half in waves 8, 10, 12, and 14; the other in waves 9, 11, and 13).

**SUPPLEMENTARY FIGURES**

**Supplementary Figure 1.** Flowchart depicting participant selection for each survey in the harmonized Gateway to Global Aging (G2A) data. Participants with at least one HbA1c measure were included. For our first analysis we excluded participants with diabetes and included those with only one HbA1c measure at baseline. For the second analysis we included participants with ≥2 HbA1c measures over time irrespective of glycemic status at baseline.

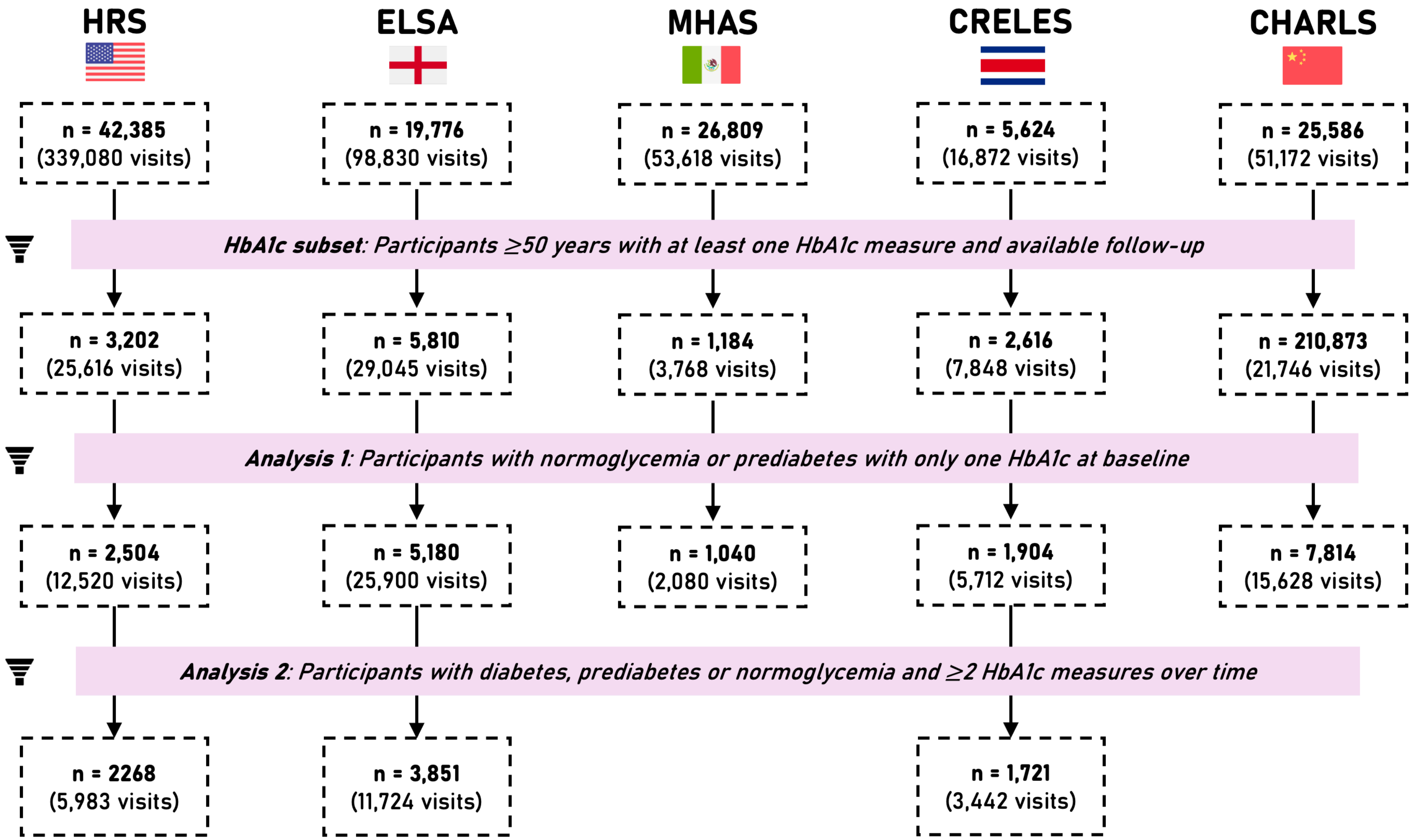

**Supplementary Figure 2.** Fixed effects Poisson regression models assessing the risk of progression and incidence of deficits in ADL, IADL, FRAIL, Fried frailty phenotype and number of comorbidities associated to a single diagnosis of prediabetes at baseline compared to normoglycemic individuals at follow-up waves t1-t4. For progression models we assessed participants with deficits at baseline, and for incidence models we assessed participants without deficits in each respective category. Models were adjusted for age, sex and cohort (**A**), and additionally for smoking, alcohol intake and BMI (**B**).

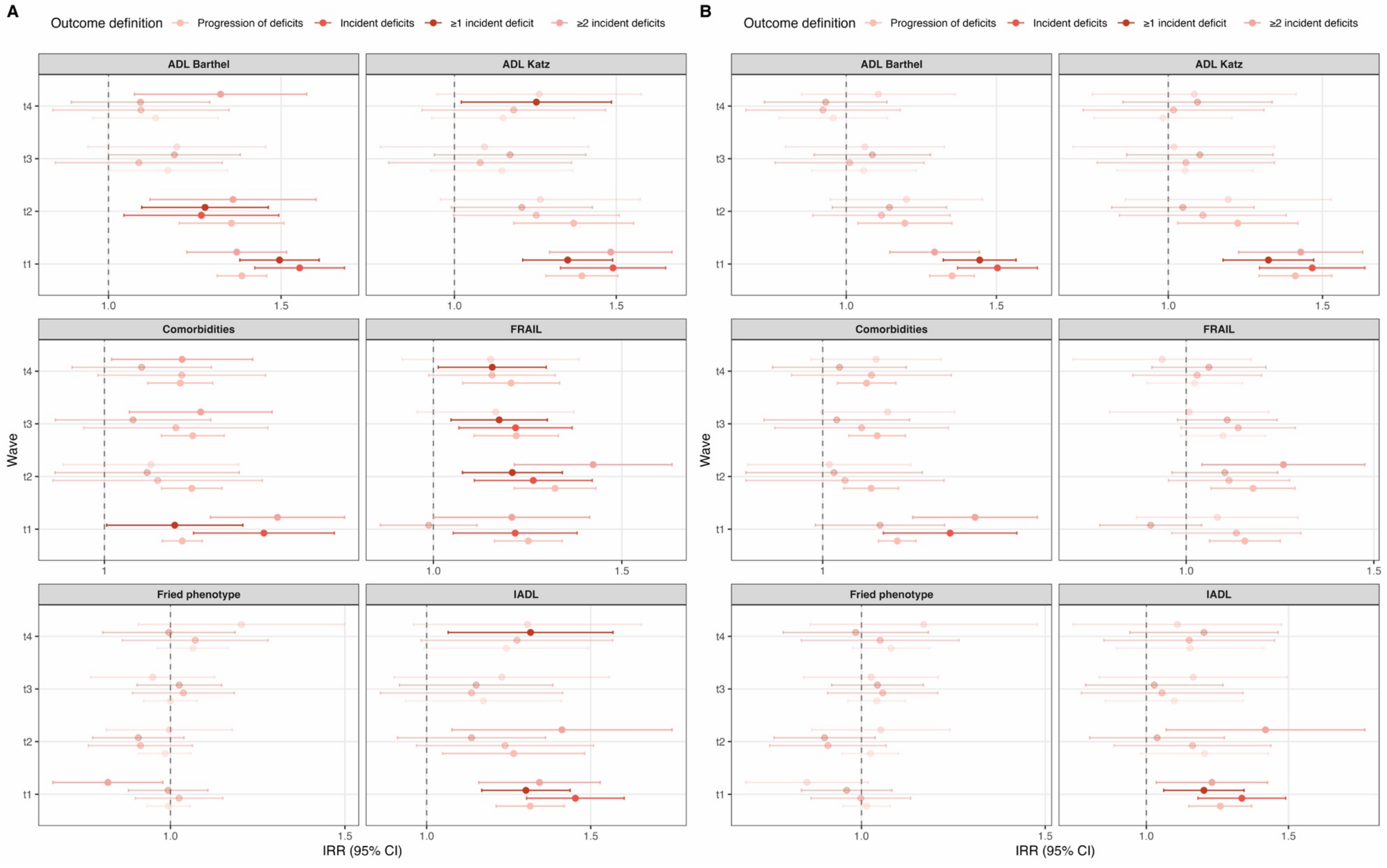

**Supplementary Figure 3.** Predicted frailty index derived from mixed effects linear regression models assessing the risk of progression and incidence of deficits in ADL, IADL, FRAIL, Fried frailty phenotype and number of comorbidities associated to a single diagnosis of prediabetes at baseline compared to normoglycemic individuals at follow-up waves t1-t4. For progression models we assessed participants with deficits at baseline, and for incidence models we assessed participants without deficits in each respective category.

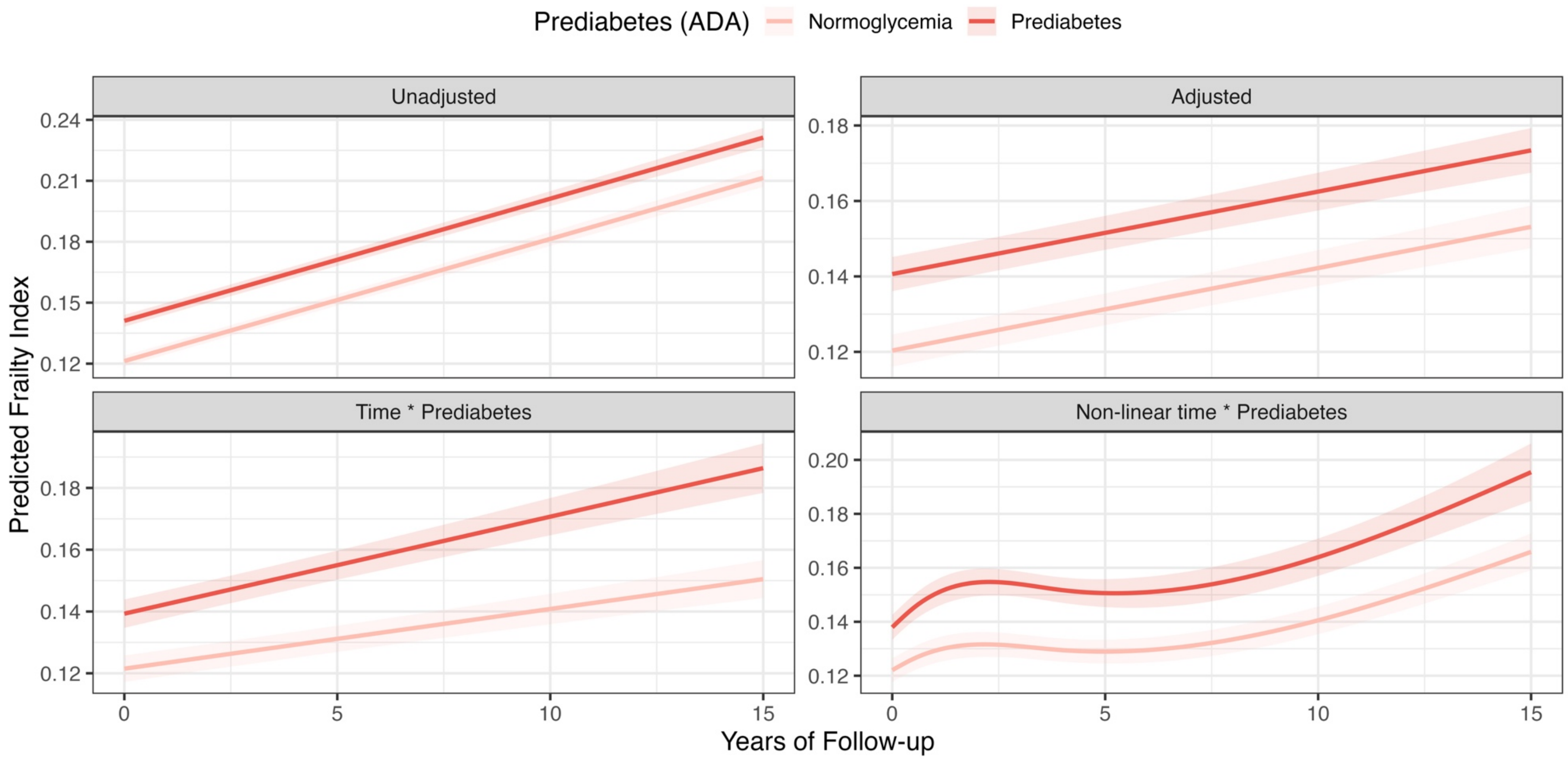

**SUPPLEMENTARY TABLES**

**Supplementary Table 1.** Included waves across each G2A study and relative time mapping.

***Abbreviations:*** *MHAS, Mexican Health and Aging Study; CRELES, Costa Rican Longevity and Healthy Aging Study; ELSA, English Longitudinal Study of Ageing; CHARLS, China Health and Retirement Longitudinal Study; HRS, Health and Retirement Study.*

| Study (Country) | Included Waves | Baseline (t=0) and Relative Time Mapping | Wave Dates |
|---|---|---|---|
| MHAS (Mexico) | Waves 03 and 04 | Baseline: Wave 03 (time=0) Follow-up: Wave 04 (time=1) | 2012 (Wave 03), 2015 (Wave 04) |
| HRS (United States) | Waves 08 to 13 | Baseline: Wave 08 (time=0) Follow-up: Wave 09 (time=1), Wave 10 (time=2), Wave 11 (time=3), Wave 12 (time=4), Wave 13 (time=5) | 2006 (Wave 08), 2008 (Wave 09), 2010 (Wave 10), 2012 (Wave 11), 2014 (Wave 12), 2016 (Wave 13) |
| CHARLS (China) | Waves 03 and 04 | Baseline: Wave 03 (time=0) Follow-up: Wave 04 (time=1) | 2015 (Wave 03), 2018 (Wave 04) |
| ELSA (England) | Waves 02, 04, 06, 08 and 09 | Baseline: Wave 02 (time=0) Follow-up: Wave 04 (time=1), Wave 06 (time=2), Wave 08 (time=3), Wave 09 (time=4) | 2004–2005 (Wave 02), 2008–2009 (Wave 04), 2012–2013 (Wave 06), 2016–2017 (Wave 08), 2018–2019 (Wave 09) |
| CRELES (Costa Rica) | Waves 01 and 02 | Baseline: Wave 01 (time=0) Follow-up: Wave 02 (time=1) | 2005 (Wave 01), 2007 (Wave 02) |

**Supplementary Table 2.** Follow-up summary for all included participants across all relative time points, number of observation, deaths and included studies.

***Abbreviations:*** *IQR, interquartile range; MHAS, Mexican Health and Aging Study; CRELES, Costa Rican Longevity and Healthy Aging Study; ELSA, English Longitudinal Study of Ageing; CHARLS, China Health and Retirement Longitudinal Study; HRS, Health and Retirement Study.*

| Time Point | Median Follow-up (years) | IQR (Interquartile Range) | Observations (n) | Deaths (n) | Participating Studies |
|---|---|---|---|---|---|
| 0 | 0.0 | (0.0 – 0.0) | 24,382 | 0 | HRS, ELSA, MHAS, CHARLS, CRELES |
| 1 | 3.0 | (2.9 – 3.7) | 20,967 | 1246 | HRS, ELSA, MHAS, CHARLS, CRELES |
| 2 | 7.6 | (4.3 – 7.8) | 6,576 | 863 | HRS, ELSA |
| 3 | 11.5 | (6.1 – 11.9) | 5,465 | 196 | HRS, ELSA |
| 4 | 13.6 | (7.9 – 14.0) | 4,800 | 225 | HRS, ELSA |

**Supplementary Table 3.** Variable harmonization across all included G2A cohorts.

***Abbreviations:*** *ADL, Activities of daily living; IADL, instrumental activities of daily living; FRAIL scale, F - Fatigue R - Resistance A - Ambulation I - Illnesses L - Loss of weight; BMI, Body-mass index; CESD, Center for Epidemiologic Studies Depression Scale; MHAS, Mexican Health and Aging Study; CRELES, Costa Rican Longevity and Healthy Aging Study; ELSA, English Longitudinal Study of Ageing; CHARLS, China Health and Retirement Longitudinal Study; HRS, Health and Retirement Study.*

| Variable / Domain | MHAS | CRELES | ELSA | CHARLS | HRS |
|---|---|---|---|---|---|
| **ID** | UNHHIDNP: Unique Person Identifier (HH ID + Person Number) | idsujeto | IDAUNIQ | ID | hhidpn: Household ID + Person ID |
| **Prediabetes by HbA1c – Baseline measurement** | reshg2_12 – wave 3 (2012) | hbac1 – wave 1 (2005) | bl_hbalc – wave 2 (2004–2005) | hba1c – wave 3 (2015) | hba1c – wave 8 (2006) |
| **Prediabetes by HbA1c – Subsequent measurements** | | hbac1 – wave 2 (2007) | hba1c – wave 4 (2008–2009); hba1c – wave 6 (2012–2013); hba1c – wave 8 (2016–2017); hba1c – wave 9 (2018–2019) | | hba1c – wave 9 (2008); hba1c – wave 10 (2010); hba1c – wave 11 (2012); hba1c – wave 12 (2014); hba1c – wave 13 (2016) |
| **Progression to diabetes** | diabe: Has had diabetes | diabe: Has had diabetes | diabe: Has had diabetes | diabe: Has had diabetes | diabe: Diabetes |
| **Activities of Daily Living (ADL) – Katz** | dressa: Difficulty dressing; batha: Difficulty bathing; beda: Difficulty getting out of bed; toilta: Difficulty using toilet; eata: | batha: Difficulty bathing; toilta: Difficulty using toilet; beda: Difficulty bed; eata: Difficulty eating | dressa: Difficulty dressing; beda: Difficulty getting out of bed; batha: Difficulty bathing; toilta: Difficulty | dressa: Difficulty dressing; batha: Difficulty bathing or showering; eata: Difficulty eating; beda: Difficulty getting in/out of bed; | dressa: Difficulty dressing; batha: Difficulty bathing; eata: Difficulty eating; beda: Difficulty getting |

| Variable / Domain | MHAS | CRELES | ELSA | CHARLS | HRS |
|---|---|---|---|---|---|
| | Difficulty eating or cutting food | | using toilet; eata: Difficulty eating | toilta: Difficulty using toilet | in/out of bed; toilta: Difficulty using toilet |
| **Activities of Daily Living (ADL) – Barthel** | dressa: Difficulty dressing; batha: Difficulty bathing; beda: Difficulty getting out of bed/need help; toilta: Difficulty using toilet; eata: Difficulty eating or cutting food; clim1a: Difficulty climbing 1 flight of stairs; walk1a: Difficulty walking 1 block (100 m) | batha: Difficulty bathing; toilta: Difficulty using toilet; beda: Difficulty bed; eata: Difficulty eating; climsa: Difficulty climbing stairs; walksa: Difficulty walking short distances | dressa: Difficulty dressing; beda: Difficulty getting out of bed; batha: Difficulty bathing; toilta: Difficulty using toilet; eata: Difficulty eating; clim1a: Difficulty climbing one flight of stairs; walk100a: Difficulty walking 100 yards | dressa: Difficulty dressing; batha: Difficulty bathing or showering; eata: Difficulty eating; beda: Difficulty getting in/out of bed; toilta: Difficulty using toilet; climsa: Difficulty climbing several flights of stairs; walk100a: Walking 100 meters | dressa: Difficulty dressing; batha: Difficulty bathing; eata: Difficulty eating; beda: Difficulty getting in/out of bed; toilta: Difficulty using toilet; walk1a: Difficulty walking 1 block; clim1a: Difficulty climbing 1 flight of stairs |
| **Instrumental Activities of Daily Living (IADL)** | mealsa: Difficulty preparing hot meal; moneya: Difficulty managing money; shopa: Difficulty shopping; medsa: Difficulty taking medications | mealsa: Difficulty preparing a meal; moneya: Difficulty managing money; shopa: Difficulty shopping; medsa: Difficulty taking medications | mealsa: Difficulty preparing hot meal; moneya: Difficulty managing money; shopa: Difficulty shopping for groceries; medsa: Difficulty taking medications | mealsa: Difficulty preparing hot meal; moneya: Difficulty managing money; shopa: Difficulty shopping for groceries; medsa: Difficulty taking medications | mealsa: Difficulty preparing a meal; moneya: Difficulty managing money; shopa: Difficulty shopping for groceries; medsa: Difficulty taking medications |
| **FRAIL Scale – Fatigue** | effort: Everything I did was an effort | c87: Severe fatigue | effort: Everything I did was an effort | effort: Everything I did was an effort | effort: Everything I did was an effort |
| **FRAIL Scale – Resistance** | clim1a: Difficulty climbing 1 flight of stairs | climsa: Difficulty climbing stairs | clim1a: Difficulty climbing one flight of stairs | climsa: Difficulty climbing several flights of stairs | clim1a: Difficulty climbing 1 flight of stairs |

| Variable / Domain | MHAS | CRELES | ELSA | CHARLS | HRS |
|---|---|---|---|---|---|
| **FRAIL Scale – Ambulation** | walk1a: Difficulty walking 1 block (100 m) | walksa: Difficulty walking short distances | walk100a: Difficulty walking 100 yards | walk100a: Walking 100 meters | walk1a: Difficulty walking 1 block |
| **FRAIL Scale – Weight loss** | c64_12: Weight change vs 2 years ago; mweight/weight | ev23: Lost weight; weight/fweight | mweight: Weight in kg | mweight: Measured weight | mweight (measured); weight (self-reported) |
| **FRAIL Scale – Comorbidities (>5)** | hibpe; hrtatte; stroke; arthre; cancre; respe | hibpe; hearte; stroke; arthre; cancre; lunge | hibpe; hearte; stroke; arthre; cancre; lunge | hibpe; hearte; stroke; arthre; cancre; lunge | hibpe; hearte; stroke; arthre; cancre (excl. skin); lunge |
| **Fried Phenotype – Fatigue** | effort: Everything I did was an effort | c87: Severe fatigue | effort: Everything I did was an effort | effort: Everything I did was an effort | effort: Everything I did was an effort |
| **Fried Phenotype – Weight loss** | c64_12: Weight change vs 2 years ago; mweight/weight | ev23: Lost weight; weight/fweight | mweight: Weight in kg | mweight: Measured weight | mweight (measured); weight (self-reported) |
| **Fried Phenotype – Weakness (grip strength)** | rgrip/lgrip: Max grip strength right/left (kg) – measured maximum | k13a/k14a: Dynamometer right/left | rgrip/lgrip: Max grip strength right/left (kg) – measured maximum | rgrip/lgrip: Max grip strength right/left (kg) – measured maximum | rgrip/lgrip: Max grip strength right/left (kg) – measured maximum |
| **Fried Phenotype – Slow gait** | wspeed1/wspeed2: Walking speed (seconds) | l6b: Walk Sec | wspeed: Average gait speed | wspeed: Walking speed (seconds) | wspeed: Average walking time |
| **Fried Phenotype – Low physical activity** | vigact: Vigorous physical activity ≥3 times/week | vigact: Vigorous physical activity ≥3 times/week | vgactx_e: Frequency of vigorous sport | vgact_c: Days per week vigorous physical activity | vgactx: Frequency of vigorous sport |

| Variable / Domain | MHAS | CRELES | ELSA | CHARLS | HRS |
|---|---|---|---|---|---|
| **Frailty Index (Deficit Accumulation)** | Proportion of 31 deficits (9 domains): ADL 5; IADL 4; CESD 5; Mobility 5; Self-reported health 3; Polypharmacy 9; Comorbidities 6; Falls 1; BMI 1 | Proportion of 26 deficits (8 domains): ADL 4; IADL 4; Mobility 3; Self-reported health 1; Polypharmacy 1; Comorbidities 6; Falls 1; BMI 1 | Proportion of 31 deficits (9 domains): ADL 5; IADL 4; CESD 5; Mobility 5; Self-reported health 3; Polypharmacy 9; Comorbidities 6; Falls 1; BMI 1 | Proportion of 31 deficits (9 domains): ADL 5; IADL 4; CESD 5; Mobility 5; Self-reported health 2; Polypharmacy 10; Comorbidities 6; Falls 1; BMI 1 | Proportion of 31 deficits (9 domains): ADL 5; IADL 4; CESD 5; Mobility 5; Self-reported health 3; Polypharmacy 11; Comorbidities 6; Falls 1; BMI 2 |

**Supplementary Table 4.** Construction of the frailty index across all included G2A cohorts. ***Abbreviations:*** *ADL, Activities of daily living; IADL, instrumental activities of daily living; FRAIL scale, F - Fatigue R - Resistance A - Ambulation I - Illnesses L - Loss of weight; BMI, Body-mass index; CESD, Center for Epidemiologic Studies Depression Scale; MHAS, Mexican Health and Aging Study; CRELES, Costa Rican Longevity and Healthy Aging Study; ELSA, English Longitudinal Study of Ageing; CHARLS, China Health and Retirement Longitudinal Study; HRS, Health and Retirement Study.*

| Variable / Domain | MHAS | CRELES | ELSA | CHARLS | HRS |
|---|---|---|---|---|---|
| Frailty Index (Deficit Accumulation) | Proportion of deficits present (31 items across 9 domains) | Proportion of deficits present (26 items across 9 domains) | Proportion of deficits present (31 items across 9 domains) | Proportion of deficits present (31 items across 9 domains) | Proportion of deficits present (31 items across 9 domains) |
| ADL | 5 items: dressa; batha; beda; toilta; eata | 4 items: batha; toilta; beda; eata (vestirse not available) | 5 items: dressa; beda; batha; toilta; eata | 5 items: dressa; batha; eata; beda; toilta | 5 items: dressa; batha; eata; beda; toilta |
| IADL | 4 items: mealsa; moneya; shopa; medsa | 4 items: mealsa; moneya; shopa; medsa | 4 items: mealsa; moneya; shopa; medsa | 4 items: mealsa; moneya; shopa; medsa | 4 items: mealsa; moneya; shopa; medsa |
| CESD (depressive symptoms) | 5 items: depres; effort; sleepr; whappy; flone | 1 item: c116 Felt with energy (No) | 5 items: depres; effort; sleepr; whappy; flone | 5 items: depres; effort; sleepr; whappy; flone | 5 items: depres; effort; sleepr; whappy; flone |
| Mobility / Physical function | 5 items: walk1a; chaira; clim1a; stoopa; armsa | 5 items: walksa; k2 Ability to stand up; climsa; d19 Difficulty cutting toenails; armsa | 5 items: walk100a; chaira; clim1a; stoopa; armsa | 5 items: walk100m; chaira; climsa; stoopa; armsa | 5 items: walk1a; chaira; clim1a; stoopa; armsa |
| Self-reported health | 3 items: shlt (poor); hearing; slfmem | 3 items: shlt; c64 Hearing; c113 Memory problems | 3 items: shlt; hearing; slfmem | 2 items: shlt; slfmem | 3 items: shlt; hearing; slfmem |
| Polypharmacy | ≥5 = 1 item: rxhibp; rxresp; rxhrtat; | ≥5 = 1 item: am31 (number of medications) | ≥5 = 1 item: rxhibp; rxlung; rxbldthn; trcancr; rxasthma; | ≥5 = 1 item: rxhibp; rxheart; cncrmeds; rxstrok; rxarthr; | ≥5 = 1 item: rxhibp; rxstrok; rxangina; rxchf; rxarthr; rxlung; |

| Variable / Domain | MHAS | CRELES | ELSA | CHARLS | HRS |
|---|---|---|---|---|---|
| | cncrmeds; rxstrok; rxarthr | | rxhrtat; rxosteo; rxdepres; rxhchol | rxdyslip; rxliver; rxkidney; rxdigest; rxmemry | rxpsych; cncrmeds; rxhrtat; rxheart; rxmemry |
| Comorbidities | 6 items: hibpe; hrtatte; stroke; arthre; cancre; respe | 6 items: hibpe; hearte; stroke; arthre; cancre; lunge | 6 items: hibpe; hearte; stroke; arthre; cancre; lunge | 6 items: hibpe; hearte; stroke; arthre; cancre; lunge | 6 items: hibpe; hearte; stroke; arthre; cancre (excl. skin); lunge |
| Falls | 1 item: fall (last 2 years) | 1 item: c49 – c49m (fall) | 1 item: fall | 1 item: da023 (fallen) | 1 item: fall (last 2 years) |
| BMI | 1 item: mbmi (measured) or <18.5; bmi (self-reported) <18.5 | 1 item: bmi <18.5 | 1 item: mbmi (measured) <18.5 | 1 item: mbmi (measured) <18.5 | 1 item: mbmi (measured) or <18.5; bmi (self-reported) <18.5 |

**Supplementary Table 5.** Age- and sex-standardized incidence of new deficits over follow-up in participants without diabetes and without any baseline deficits in any of their respective categories amongst included participants in G2A cohorts stratified by glycemic status in prediabetes and diabetes. ***Abbreviations:*** *ADL, Activities of daily living; IADL, instrumental activities of daily living; FRAIL scale, F - Fatigue R - Resistance A - Ambulation I - Illnesses L - Loss of weight; IRR, Incidence rate ratio; 95%CI, 95% confidence interval*.

| Incident deficit | Status | Participants at risk | Person-years | Incident category | Events | Age- and sex-standardized incidence | Lower 95%CI | Upper 95%CI |
|---|---|---|---|---|---|---|---|---|
| ADL-Katz | Normoglycemia | 9,151 | 63,985.42 | ≥1 deficit | 1,445 | 25.20551 | 23.89021 | 26.58032 |
| | | | | ≥2 deficits | 675 | 12.18057 | 11.25736 | 13.16477 |
| | Prediabetes | 6,404 | 30,322.08 | ≥1 deficit | 1,001 | 34.77301 | 32.61736 | 37.04649 |
| | | | | ≥2 deficits | 457 | 16.16998 | 14.69593 | 17.76422 |
| ADL-Barthel | Normoglycemia | 7,315 | 56,448.00 | ≥1 deficit | 1,960 | 39.50647 | 37.68895 | 41.40068 |
| | | | | ≥2 deficits | 822 | 18.01992 | 16.74816 | 19.37253 |
| | Prediabetes | 4,590 | 24,220.08 | ≥1 deficit | 1,427 | 61.13364 | 57.92469 | 64.50207 |
| | | | | ≥2 deficits | 519 | 23.85161 | 21.78219 | 26.08944 |
| IADL | Normoglycemia | 9,206 | 66,379.75 | ≥1 deficit | 1,444 | 25.34405 | 24.01111 | 26.73910 |
| | | | | ≥2 deficits | 660 | 12.35030 | 11.39888 | 13.36629 |
| | Prediabetes | 6,455 | 31,586.75 | ≥1 deficit | 1,029 | 34.28112 | 32.18429 | 36.49115 |
| | | | | ≥2 deficits | 454 | 15.72381 | 14.28432 | 17.28024 |
| FRAIL | Normoglycemia | 5,682 | 47,259.00 | ≥1 deficit | 2,437 | 55.72016 | 53.41764 | 58.11273 |
| | | | | ≥2 deficits | 826 | 21.21295 | 19.71900 | 22.80358 |

| Incident deficit | Status | Participants at risk | Person-years | Incident category | Events | Age- and sex-standardized incidence | Lower 95%CI | Upper 95%CI |
|---|---|---|---|---|---|---|---|---|
| | Prediabetes | 3,027 | 18,247.08 | ≥1 deficit | 908 | 51.88689 | 48.48377 | 55.50343 |
| | | | | ≥2 deficits | 319 | 19.01104 | 16.92426 | 21.32010 |
| Fried deficits | Normoglycemia | 4,237 | 29,288.42 | ≥1 deficit | 1,873 | 66.48375 | 63.40198 | 69.68988 |
| | | | | ≥2 deficits | 724 | 25.47773 | 23.58937 | 27.49101 |
| | Prediabetes | 2,799 | 13,403.00 | ≥1 deficit | 586 | 46.09844 | 42.35545 | 50.13056 |
| | | | | ≥2 deficits | 238 | 18.80017 | 16.43554 | 21.45887 |
| Number of comorbidities | Normoglycemia | 3,825 | 27,363.00 | ≥1 deficit | 1,048 | 40.49963 | 37.86017 | 43.32974 |
| | | | | ≥2 deficits | 438 | 16.91527 | 15.22785 | 18.79683 |
| | Prediabetes | 2,391 | 11,470.75 | ≥1 deficit | 612 | 53.91194 | 49.47684 | 58.76806 |
| | | | | ≥2 deficits | 241 | 23.21027 | 20.15974 | 26.71472 |

**Supplementary Table 5.** Mixed effects linear regression models assessing the impact of a single diagnosis of prediabetes at baseline on changes of the frailty index over time compared to normoglycemic individuals amongst participants without diabetes from included G2A cohorts. Models were adjusted for time-varying age, sex and cohort of origin, and had a random slope for time and a random intercept to account for repeated measures over time.

| Model | Term | β-coefficient (95% CI) | p-value |
|---|---|---|---|
| Unadjusted model | Prediabetes (baseline) | 0.020 (0.016, 0.024) | <0.001 |
| Fully adjusted model | Prediabetes (baseline) | 0.020 (0.017, 0.024) | <0.001 |
| Interaction model | Prediabetes (baseline) | 0.018 (0.014, 0.022) | <0.001 |
| | Prediabetes × Follow-up years | 0.001 (0.001, 0.002) | <0.001 |
| Interaction with non-linear time | Prediabetes (baseline) | 0.016 (0.012, 0.020) | <0.001 |
| | Prediabetes × ns(Follow-up years, df = 3)1 | -0.001 (-0.013, 0.011) | 0.833 |
| | Prediabetes × ns(Follow-up years, df = 3)2 | 0.016 (0.008, 0.024) | <0.001 |
| | Prediabetes × (ns(Follow-up years, df = 3)3 | 0.010 (-0.002, 0.021) | 0.0938 |

**Supplementary Table 6.** Glycemic transitions over time amongst participants with ≥2 HbA1c measurements over time in HRS, ELSA and CRELES. To assess transitions, denominators only consider participants from the previous follow-up if they had available HbA1c measures at follow-up.

| Wave transitions | Prior glycemic status | Glycemic status at follow-uo | Number of participants | Proportion within prior status |
|---|---|---|---|---|
| From wave 1 to wave 2 N=7,840 | Normoglycemia | Normoglycemia | 2,587 | 51.5% |
| | | Prediabetes | 1,868 | 37.2% |
| | | Diabetes | 567 | 11.3% |
| | Prediabetes | Normoglycemia | 348 | 20.8% |
| | | Prediabetes | 916 | 54.9% |
| | | Diabetes | 406 | 24.3% |
| | Diabetes | Diabetes | 1,148 | 100.0% |
| From wave 2 to wave 3 N= 4,083 | Normoglycemia | Normoglycemia | 1,420 | 77.1% |
| | | Prediabetes | 390 | 21.2% |
| | | Diabetes | 31 | 1.7% |
| | Prediabetes | Normoglycemia | 551 | 34.4% |
| | | Prediabetes | 949 | 59.2% |
| | | Diabetes | 104 | 6.5% |
| | Diabetes | Diabetes | 638 | 100.0% |
| From wave 3 to wave 4 N= 1,386 | Normoglycemia | Normoglycemia | 632 | 89.6% |
| | | Prediabetes | 70 | 9.9% |
| | | Diabetes | 3 | 0.4% |
| | Prediabetes | Normoglycemia | 230 | 44.1% |
| | | Prediabetes | 268 | 51.3% |
| | | Diabetes | 24 | 4.6% |
| | Diabetes | Diabetes | 159 | 100.0% |

**Supplementary Table 7.** Generalized Estimating Equation models with a Poisson link assessing the risk of incident deficits in ADL, IADL, FRAIL, Fried frailty and number of comorbidities associated to stable prediabetes, progression to diabetes or regression to normoglycemia among participants with prediabetes from ELSA, HRS and CRELES with ≥1 HbA1c measurement over time.

***Abbreviations:*** *ADL, Activities of daily living; IADL, instrumental activities of daily living; FRAIL scale, F - Fatigue R - Resistance A - Ambulation I - Illnesses L - Loss of weight; IRR, Incidence rate ratio; 95%CI, 95% confidence interval*

***Note:*** *For incidence models we assessed participants without deficits in each respective category. Models were adjusted for age, cohort and sex.*

| Outcome | Term | IRR | 95%CI low | 95%CI high | p-value |
|---|---|---|---|---|---|
| ADL-Katz | Progression to diabetes | 0.42 | 0.20 | 0.90 | 0.025 |
| | Regression to normoglycemia | 1.71 | 0.91 | 3.20 | 0.095 |
| | Progression*Time | 1.27 | 1.09 | 1.48 | 0.002 |
| | Regression*Time | 1.01 | 0.87 | 1.17 | 0.892 |
| ADL-Bathel | Progression to diabetes | 0.64 | 0.32 | 1.27 | 0.202 |
| | Regression to normoglycemia | 1.88 | 1.14 | 3.09 | 0.013 |
| | Progression*Time | 1.22 | 1.06 | 1.41 | 0.005 |
| | Regression*Time | 0.98 | 0.87 | 1.10 | 0.683 |
| IADL | Progression to diabetes | 0.64 | 0.19 | 2.16 | 0.469 |
| | Regression to normoglycemia | 1.90 | 0.78 | 4.65 | 0.159 |
| | Progression*Time | 1.32 | 1.03 | 1.68 | 0.027 |
| | Regression*Time | 1.12 | 0.93 | 1.34 | 0.235 |
| FRAIL | Progression to diabetes | 0.95 | 0.52 | 1.74 | 0.867 |

| Outcome | Term | IRR | 95%CI low | 95%CI high | p-value |
|---|---|---|---|---|---|
| | Regression to normoglycemia | 2.06 | 1.42 | 3.00 | 0.000 |
| | Progression*Time | 1.02 | 0.88 | 1.18 | 0.810 |
| | Regression*Time | 0.94 | 0.87 | 1.01 | 0.102 |
| Fried | Progression to diabetes | 0.34 | 0.21 | 0.55 | 0.000 |
| | Regression to normoglycemia | 1.58 | 1.24 | 2.01 | 0.000 |
| | Progression*Time | 1.20 | 0.95 | 1.52 | 0.126 |
| | Regression*Time | 0.97 | 0.90 | 1.04 | 0.334 |
| Number of comorbidities | Progression to diabetes | 1.00 | 0.76 | 1.30 | 0.975 |
| | Regression to normoglycemia | 1.04 | 0.87 | 1.25 | 0.679 |
| | Progression*Time | 1.08 | 0.99 | 1.19 | 0.096 |
| | Regression*Time | 1.02 | 0.98 | 1.07 | 0.350 |

**Supplementary Table 8.** Generalized Estimating Equation models with a Poisson link assessing the risk of incident deficits in ADL, IADL, FRAIL, Fried frailty and number of comorbidities associated to stable prediabetes, progression to diabetes or regression to normoglycemia among participants with prediabetes from ELSA, HRS and CRELES with ≥1 HbA1c measurement over time **after additional adjustment by lifestyle variables and body-mass index**.

***Abbreviations:*** *ADL, Activities of daily living; IADL, instrumental activities of daily living; FRAIL scale, F - Fatigue R - Resistance A - Ambulation I - Illnesses L - Loss of weight; IRR, Incidence rate ratio; 95%CI, 95% confidence interval*

***Note:*** *For incidence models we assessed participants without deficits in each respective category. Models were adjusted for age, cohort, sex, smoking, drinking, and BMI.*

| Outcome | Term | IRR | 95%CI low | 95%CI high | p-value |
|---|---|---|---|---|---|
| ADL-Katz | Progression to diabetes | 0.15 | 0.06 | 0.38 | 0.000 |
| | Regression to normoglycemia | 1.33 | 0.72 | 2.46 | 0.368 |
| | Progression*Time | 1.45 | 1.22 | 1.72 | 0.000 |
| | Regression*Time | 1.03 | 0.90 | 1.18 | 0.621 |
| ADL-Bathel | Progression to diabetes | 0.30 | 0.12 | 0.73 | 0.009 |
| | Regression to normoglycemia | 1.49 | 0.88 | 2.54 | 0.141 |
| | Progression*Time | 1.34 | 1.12 | 1.59 | 0.001 |
| | Regression*Time | 1.01 | 0.90 | 1.14 | 0.818 |
| IADL | Progression to diabetes | 0.24 | 0.06 | 0.99 | 0.048 |
| | Regression to normoglycemia | 2.02 | 0.78 | 5.23 | 0.146 |
| | Progression*Time | 1.45 | 1.14 | 1.85 | 0.002 |
| | Regression*Time | 1.08 | 0.89 | 1.30 | 0.442 |
| FRAIL | Progression to diabetes | 0.71 | 0.35 | 1.42 | 0.329 |

| Outcome | Term | IRR | 95%CI low | 95%CI high | p-value |
|---|---|---|---|---|---|
| | Regression to normoglycemia | 2.03 | 1.36 | 3.04 | 0.001 |
| | Progression*Time | 1.06 | 0.90 | 1.25 | 0.483 |
| | Regression*Time | 0.95 | 0.87 | 1.03 | 0.208 |
| Fried | Progression to diabetes | 0.37 | 0.22 | 0.62 | 0.000 |
| | Regression to normoglycemia | 1.59 | 1.25 | 2.01 | 0.000 |
| | Progression*Time | 1.22 | 0.97 | 1.53 | 0.093 |
| | Regression*Time | 0.96 | 0.90 | 1.03 | 0.225 |
| Number of comorbidities | Progression to diabetes | 0.82 | 0.63 | 1.07 | 0.149 |
| | Regression to normoglycemia | 1.05 | 0.88 | 1.26 | 0.557 |
| | Progression*Time | 1.10 | 1.00 | 1.22 | 0.051 |
| | Regression*Time | 1.02 | 0.98 | 1.07 | 0.329 |
